# Distinct genomic signatures and modifiable risk factors underly the comorbidity between major depressive disorder and cardiovascular disease

**DOI:** 10.1101/2023.09.01.23294931

**Authors:** Jacob Bergstedt, Joëlle A. Pasman, Ziyan Ma, Arvid Harder, Shuyang Yao, Nadine Parker, Jorien L. Treur, Dirk J.A. Smit, Oleksandr Frei, Alexey Shadrin, Joeri J. Meijsen, Qing Shen, Sara Hägg, Per Tornvall, Alfonso Buil, Thomas Werge, Jens Hjerling-Leffler, Thomas D. Als, Anders D. Børglum, Cathryn M. Lewis, Andrew M. McIntosh, Unnur A. Valdimarsdóttir, Ole A. Andreassen, Patrick F. Sullivan, Yi Lu, Fang Fang

## Abstract

Major depressive disorder (MDD) and cardiovascular disease (CVD) are often comorbid, resulting in excess morbidity and mortality. Using genomic data, this study elucidates biological mechanisms, key risk factors, and causal pathways underlying their comorbidity. We show that CVDs share a large proportion of their genetic risk factors with MDD. Multivariate genome-wide association analysis of the shared genetic liability between MDD and atherosclerotic CVD (ASCVD) revealed seven novel loci and distinct patterns of tissue and brain cell-type enrichments, suggesting a role for the thalamus. Part of the genetic overlap was explained by shared inflammatory, metabolic, and psychosocial/lifestyle risk factors. Finally, we found support for causal effects of genetic liability to MDD on CVD risk, but not from most CVDs to MDD, and demonstrated that the causal effects were partly explained by metabolic and psychosocial/lifestyle factors. The distinct signature of MDD-ASCVD comorbidity aligns with the idea of an immunometabolic sub-type of MDD more strongly associated with CVD than overall MDD. In summary, we identify plausible biological mechanisms underlying MDD-CVD comorbidity, as well as key modifiable risk factors for prevention of CVD in individuals with MDD.

## Main

Major depressive disorder (MDD) and cardiovascular disease (CVD) are highly comorbid^1,2^. Several mechanisms might explain the observed comorbidity^2^. One explanation is that genetic risk factors for MDD and CVDs overlap ^3,4^. While observed genome-wide genetic correlations between MDD and CVD are modest^2–4^, this may be because local genetic correlations of opposing directions attenuate correlations on the genome-wide level, leading to an underestimation of the genetic overlap^5^. The large polygenicity of MDD^6^ might also mask subtypes with stronger genetic relationships to CVD.

The observed MDD-CVD comorbidity could also be due to non-genetic factors^7^. Cardiovascular risk factors like high systolic blood pressure, high body mass index (BMI), high levels of low-density lipoprotein cholesterol, high levels of physical inactivity, presence of type 2 diabetes, and smoking have all been associated with MDD^8–10^. Moreover, accumulating data show that psychosocial/lifestyle factors associated with MDD, like low educational attainment, exposure to childhood maltreatment, loneliness, and atypical sleep patterns, are also important risk factors for CVD^11–14^.

One common mechanism underlying MDD, CVD, as well as their shared risk factors could be chronic inflammation. Atherosclerosis, the accumulation of fibrofatty lesions in the arterial wall, is the main cause of CVD^15^. The build-up of atherosclerotic plaque is a long-term inflammatory process mediated by immune components in crosstalk with arterial wall cells^16^. Many lines of evidence also support a role for inflammation in MDD^17^. Excessive or long-term psychosocial stress promote the maturation and release of inflammatory cytokines like interleukin (IL)-6, which activate the central nervous system to produce behaviors related to MDD^18^. Importantly, low-grade inflammation, defined by high C-reactive protein levels, has been observed in more than a quarter of patients with depression^19^, suggesting the presence of an inflammatory subtype of MDD^20^, which might be especially strongly associated with CVD.

The full extent of the genetic overlap between MDD and CVD has not been explored. It remains unknown if the genetic overlap is associated with specific tissues or brain cell-types, or how this overlap relates to shared risk factors such as blood pressure, psychosocial/lifestyle traits, metabolic traits, and inflammation. Moreover, causal effects linking these traits are not fully understood ^21–24^.

Here, we dissect the genetic overlap between MDD and CVD (coronary artery disease, peripheral artery disease, heart failure, stroke, and atrial fibrillation) by leveraging state-of-the-art genomic data and methods (Fig. 1). We used newly released summary statistics from a genome-wide association study (GWAS) of MDD involving 290,000 cases^6^, with substantially increased statistical power compared to previous GWASs. First, we assessed the pairwise genetic overlap between MDD and CVD on the genome-wide level, as well as on the level of local partitions of the genome and overlapping causal variants with MiXeR^25^ and LAVA^26^. Using these methods, we were able to consider the direction of correlation at each locus in the genome, providing a more granular understanding of the genetic overlap between MDD and CVD. Second, we identified genetic variants and genes that contribute to the shared liability between MDD and atherosclerotic CVD (ASCVD; coronary artery disease, peripheral artery disease, heart failure, and stroke) using Genomic Structural Equation Modeling (SEM)^27^. We mapped identified variants to brain cell-types using novel annotations based on single-cell RNA sequencing in post-mortem human brain samples^28^. Third, we assessed shared risk factors explaining the association between MDD and CVD. To do so, we evaluated the polygenic overlap between MDD and well-established risk factors. We then estimated genetic correlation between MDD and CVD adjusting for risk factors, and the genetic correlation between the shared genetic liability between MDD and ASCVD and risk factors. Finally, we used Mendelian Randomization (MR) to investigate putative causal effects and assess mediation through shared risk factors for MDD and CVD. In brief, this study leverages recent large-scale GWAS data and triangulates results from current genomic methods to elucidate etiological pathways underlying the comorbidity between MDD and CVD.

**Figure 1.**
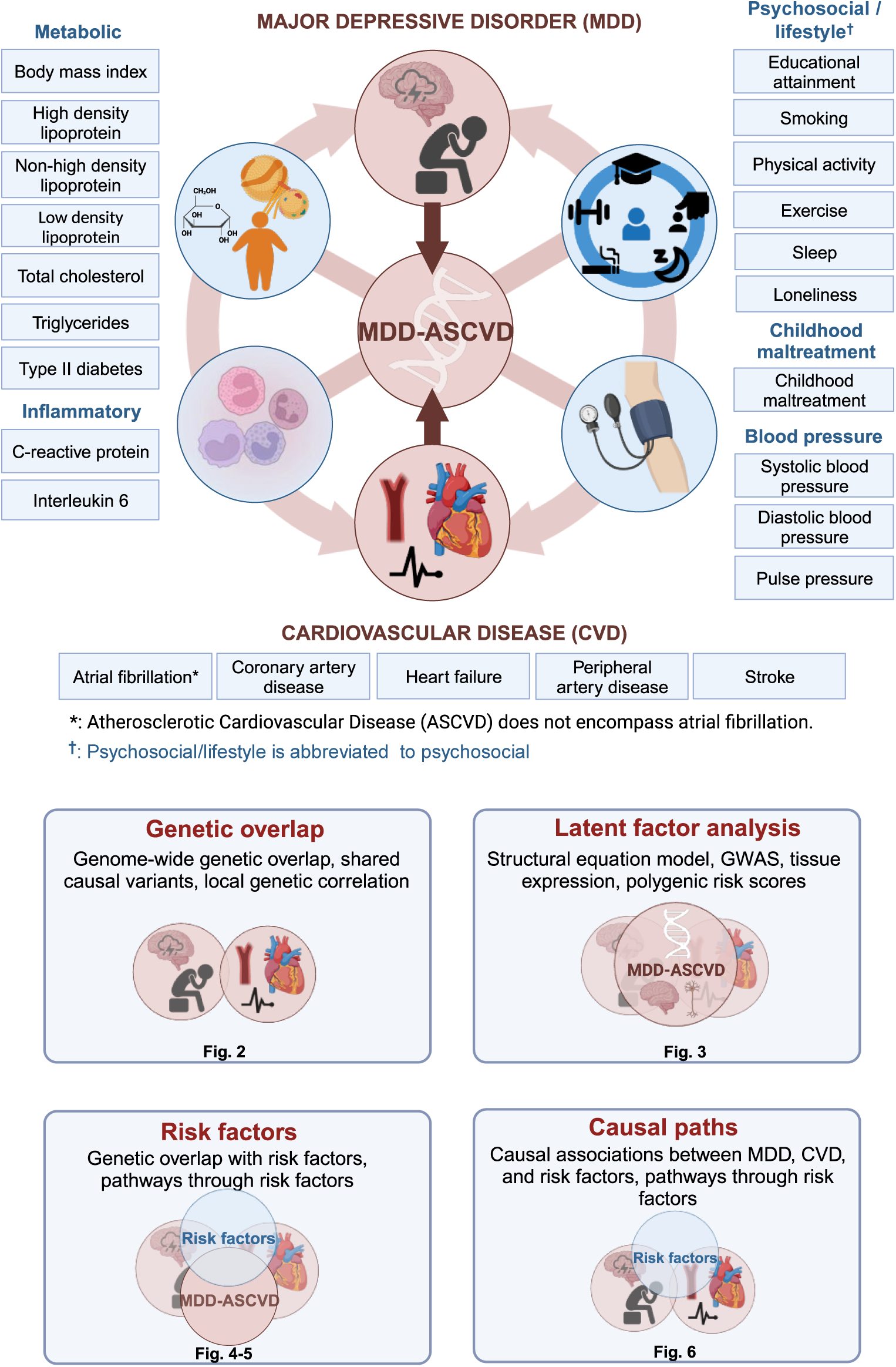
Graphical abstract illustrating the study approach. The comorbidity between MDD and CVD is investigated using genetic and causal inference methods, including assessing overlap with and mediation through shared risk factors (metabolic, psychosocial/lifestyle, inflammatory, and blood pressure; Supplementary Table 1). The risk factor group psychosocial/lifestyle is abbreviated to psychosocial. Created with BioRender.com (license agreements VG26BG3VTL, MK26BG46H3).

## Results

### Most genetic risk factors for CVD overlap with MDD

There were weak to moderate genome-wide genetic correlations between MDD and CVDs, as estimated with linkage disequilibrium score regression (LDSC^29^; Extended Data Fig. 1, Supplementary Table 2). Information on source GWASs for all traits is given in Supplementary Table 1. The strongest correlations were noted for peripheral artery disease (*r_g_*=0.30, SE=0.04, *P*=1×10^-13^), heart failure (*r_g_*=0.29, SE=0.03, *P*=1×10^-24^), and coronary artery disease (*r_g_*=0.25, SE=0.02, *P*=9×10^-45^), while smaller but statistically significant correlations were observed for stroke and atrial fibrillation (*r_g_*=0.18 and *r_g_*=0.11, *P*<1×10^-7^). ASCVDs showed strong correlations among each other, while atrial fibrillation was only moderately genetically correlated to the ASCVDs (Extended Data Fig. 2a, Supplementary Table 2). To aid interpretation of these results, we also analyzed specific MDD symptoms. Poor appetite or overeating was consistently the symptom that showed the strongest genetic correlation to the CVDs (Extended Data Fig. 1a).

We then estimated local genetic correlations between MDD and CVDs in each of 2,495 distinct genomic regions used in LAVA^26^. The distribution of local genetic correlations is displayed in Fig. 2a, with points exceeding the horizontal line representing partitions with a genome-wide significant correlation (*P_FDR_*<0.05). We found 54 significant local correlations between MDD and CVDs, 40 of which were for MDD and coronary artery disease. Most significant local correlations were positive (90%), although results were mixed for peripheral artery disease and heart failure, with 50% and 25% of the partitions, respectively, showing negative correlations with MDD (Supplementary Table 4). We also investigated local genetic correlation between MDD and the CVDs in 16 loci in the human leukocyte antigen (HLA) region (Extended Data Fig 1b). Out of 50 assessed local genetic correlations, 8 were significant, indicating that this region is a hotspot of genetic correlation between MDD and CVDs.

**Figure 2.**
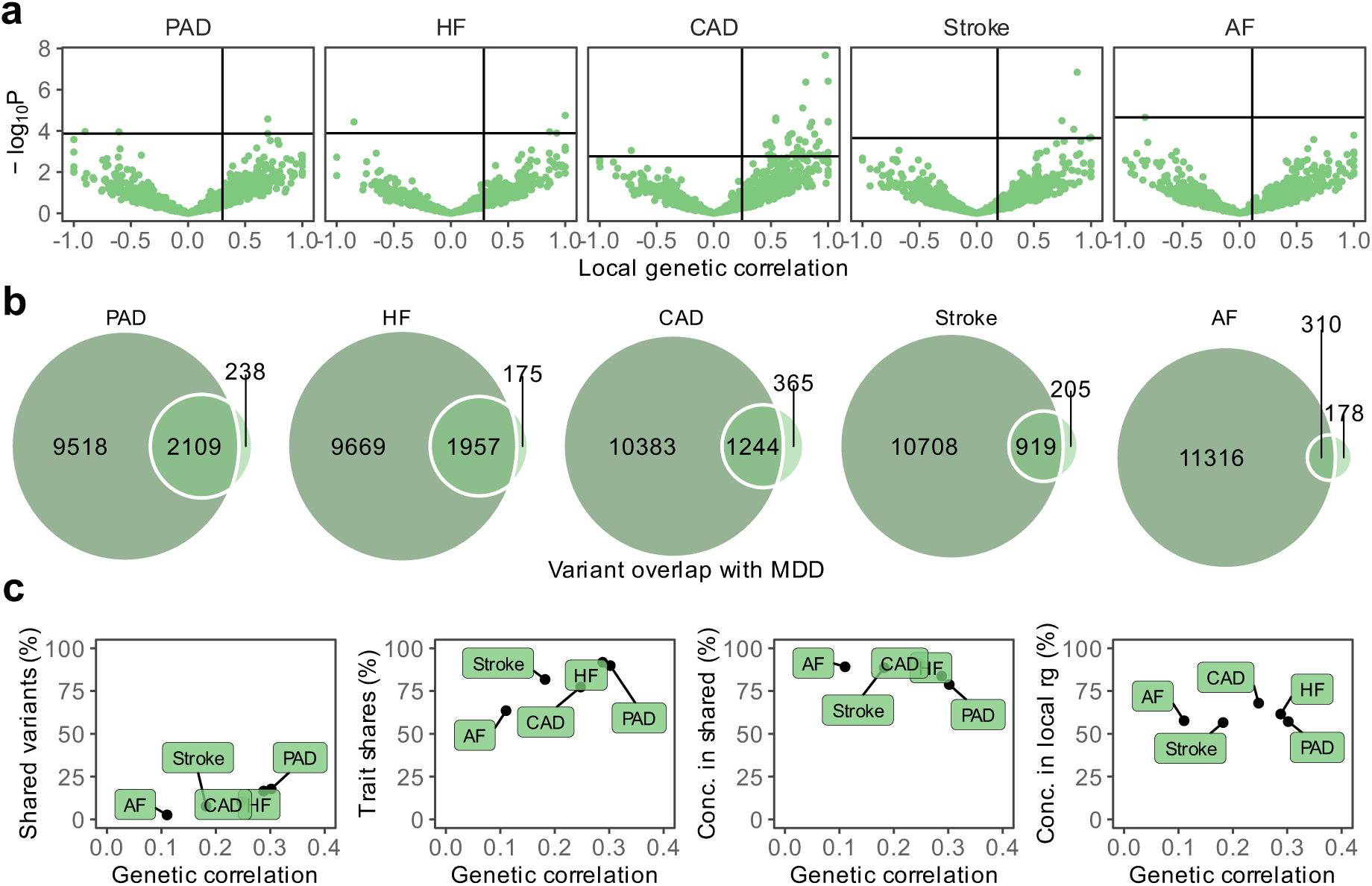
Genetic overlap between MDD and CVD beyond genome-wide genetic correlation. (**a)** Volcano plots based on LAVA results showing genomic loci (green dots) with the local genetic correlation between MDD and each of the CVDs (x-axis) and the corresponding *P*-value (y-axis). Loci exceeding the horizontal line are significant at *P*_FDR_<0.05. Multiple testing was performed separately for each trait over all considered loci (**b)** Venn diagrams based on MiXeR results showing the number of causal variants that are unique to MDD (left circle), unique to CVD (non-overlapping part of right circle), or shared between MDD and CVD (overlapping part of circles). (**c)** Genetic correlation estimated by LDSC (x-axis) against the percentage of MDD variants that are shared with the CVD trait as estimated by MiXeR (first plot), the percentage of CVD variants that are shared with MDD (second plot), and the percentage of CVD variants that are shared with MDD that have concordant effect directions (third plot). The fourth plot shows the percentage of local genetic correlations from LAVA that have concordant effect directions on the y-axis. **a-c** Sample sizes and information for underlying summary statistics GWASs are reported in Supplementary Table 1. MDD=Major Depressive Disorder, PAD=Peripheral Artery Disease, CAD=Coronary Artery Disease, AF=Atrial Fibrillation, HF=Heart Failure

Next, we investigated genetic overlap on the level of risk variants using MiXeR^25^ (Supplementary Tables 5-6). We identified more variants for MDD than for the CVDs, suggesting that MDD is more polygenic than CVD. To verify these results, we estimated polygenicity using a complementary Bayesian approach implemented in SBayesS^30^, which showed estimates that were highly correlated with those of MiXeR (Extended Data Fig. 2b, Supplementary Table 7).

Bivariate MiXeR results showed that CVDs shared a large proportion of their causal variants with MDD (from 64% in atrial fibrillation to 92% in heart failure, Fig. 2b) whereas MDD shared only a small proportion of its causal variants with CVDs (<20%). Note that for peripheral artery disease the performance metrics of the model indicate that this finding needs to be interpreted with caution (Supplementary Table 6). Both shared genetic variants and local genetic correlations exhibit strong degrees of effect direction concordance (Fig. 2c), suggesting that genetic risk variants for CVDs are strongly correlated with a genetic subcomponent of MDD.

### Shared genetic liability to MDD and CVD

To further characterize the genetic overlap, we explicitly modelled the shared liability between MDD and CVD as a higher-order latent factor using Genomic SEM. We excluded atrial fibrillation because it deteriorated the model fit (CFI=0.918, SRMR=0.072) and had the lowest factor loading (*β*=0.56, SE=0.05), so that the interpretation of the latent factor changed to representing ASCVD. The final model had an excellent fit with Comparative Fit Index CFI>0.999 and Standardized Root Mean Squared Residual SRMR=0.021. The factor loading for MDD was *β*=0.32. The loadings for the ASCVDs ranged from *β*=0.63 for stroke (SE=0.06, *P*=4.37×10^-41^) to *β*=0.85 for heart failure (SE=0.07, *P*=2.97×10^-56^) (loadings standardized on the latent factors are shown in Fig. 3a). For comparison, we also fit a latent factor for the ASCVDs alone (without MDD), which showed similar fit and parameter estimates (CFI>0.999, SRMR=0.013) (Extended Data Fig. 3a). This shows that shared genetic liability to different ASCVDs as well as to ASCVDs and MDD (to a lesser extent) can be explained by a single underlying factor.

**Figure 3.**
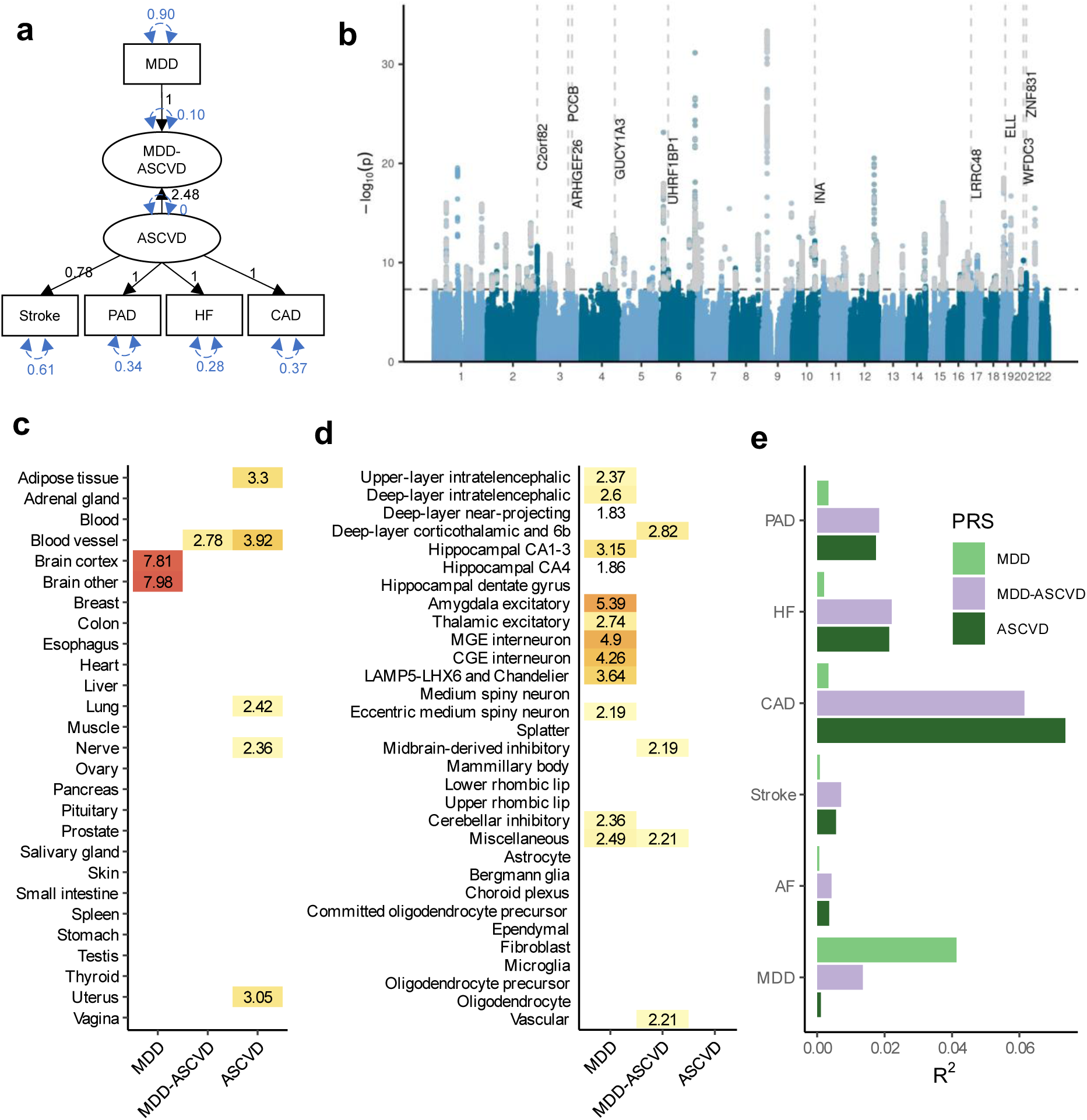
Shared genetic liability factor for MDD-ASCVD. (**a)** Latent factor model as specified in Genomic SEM with the ‘observed’ variables in rectangles and the latent variables in circles. Factor loadings (standardized on the latent factors) are given in black and variances in blue. Sample sizes for underlying GWAS summary statistics are reported in Supplementary Table 1 (**b)** Latent MDD-ASCVD factor GWAS results. The x-axis shows genomic position, and the y-axis shows statistical significance as –log_10_(*P*). Genome-wide significant SNPs that were filtered out because of significant heterogeneity Q_SNP_ are displayed in grey. The top 10 eQTL genes are displayed with dashed vertical lines indicating their position. (**c)** Enrichment results in GTEx tissues for the latent MDD-ASCVD factor, with latent ASCVD (without MDD) and MDD-only as comparison. (**d)** Enrichment results for the latent MDD-ASCVD factor, latent ASCVD, and MDD-only in brain cell types. **(e)** Proportion of variance explained in MDD and CVD phenotypes in the UKB (defined using ICD-codes listed in Supplementary Table 14) by each of 3 PRSs for the latent MDD-ASCVD factor, latent ASCVD, or MDD-only. **c, d** Enrichment is measured using significance testing in a two-sided t-test displayed as −log_10_(*P*). Only tissues with a significant association (*P*_FDR_<0.05) are shown. Multiple testing was performed over tested tissues/cell-types. MDD=Major Depressive Disorder; MDD-ASCVD=common factor for MDD and ASCVD; ASCVD=common factor for the atherosclerotic cardiovascular diseases

The GWAS on the latent MDD-ASCVD factor resulted in 205 independent genome-wide significant loci (Fig. 3b, independent at R^2^<0.1 and distance≥250kb, Supplementary Table 8). We did not observe genomic inflation, indicating that results were not strongly affected by population stratification (LDSC intercept=1.02). Almost three quarters (74.6%) of the genome-wide statistically significant SNPs showed a high Q heterogeneity, suggesting that their effects were more in line with an independent pathway than a common pathway model (see methods). Most of this heterogeneity was due to MDD, as the GWAS for latent ASCVD without MDD showed fewer genome-wide statistically significant SNPs with a high heterogeneity (30.6%; Extended Data Fig. 3b).

For the latent MDD-ASCVD factor, we filtered out variants that showed significant heterogeneity and considered only variants where the latent MDD-ASCVD factor was the best model for the follow-up analyses. We retained 72 independent loci underlying the shared genetic liability (Fig. 3b, Supplementary Table 9). The top SNP after filtering was rs11670056 in the *ELL* gene on chromosome 19, which is part of the transcription elongation factor complex and has previously been associated with a range of CVDs, blood traits, BMI, and educational attainment (enrichment in associations with other traits for significant SNPs are shown in Extended Data Fig. 4a). There were 19 top SNPs that were significant eQTLs for one or multiple genes (top 10 are annotated in Fig. 3b, full results are shown in Supplementary Table 10). Besides *ELL,* multiple genes on chromosome 10 around *INA* and *CNNM2* were identified. *INA* is involved in structural neuron regulation whereas *CNNM2* is involved in ion transportation and has previously been associated with psychiatric as well as cardiovascular traits.

From the latent MDD-ASCVD GWAS summary statistics we extracted seven novel loci that were not among the risk loci in the MDD and ASCVD GWASs that constituted the latent factor (Extended Data Fig. 4b, Supplementary Table 11). The top SNPs in these loci have not been identified in any GWAS recorded in the GWAS catalog before, but four of them have shown suggestive associations (*P*<0.05) with metabolic traits (rs11065577, rs11606884, rs2838351, and rs500571).

Using partitioned LDSC, we observed that the heritability of the latent MDD-ASCVD factor was enriched in genes with expression specific to endothelial and blood vessel tissues, which was also observed for latent ASCVD, but not for MDD (Fig. 3c, Supplementary Table 12). To gain deeper insights into brain-specific mechanisms, we leveraged high-resolution human brain single-nucleus RNAseq data^28^ and identified four human brain cell-types that exhibited enriched MDD-ASCVD heritability, including deep layer corticothalamic and 6b cells, midbrain-derived inhibitory neurons, miscellaneous neurons, and vascular cells (Fig. 3d, Supplementary Table 13). Notably, three of the four cell-types were uniquely associated with the latent MDD-ASCVD factor, displaying no enrichment for either latent ASCVD without MDD or MDD only, suggesting that the genetic variance for MDD-ASCVD comorbidity has a distinct functional signature.

To externally validate the MDD-ASCVD phenotype, we computed polygenic risk scores (PRS) based on the summary statistics for the latent MDD-ASCVD factor, as well as for MDD and latent ASCVD, and found them to significantly predict ASCVD and MDD diagnoses in UK Biobank (UKB) (all *P*<2×10^-13^; Supplementary Tables 14-15; note that source and target samples were overlapping and the R^2^ values should only be interpreted relative to one another). The PRS for latent ASCVD and the latent MDD-ASCVD factor explained similar amounts of variance in ASCVD (Fig. 3e). This is in line with the MiXeR findings (Fig. 2b), suggesting that most causal variants for ASCVD are shared with MDD. In contrast, as most causal variants for MDD are not shared with ASCVD, the PRS for the latent MDD-ASCVD factor explained less than half as much variance in MDD as the MDD PRS.

Next, we assessed genetic correlations between the latent MDD-ASCVD factor and MDD symptoms. We found that poor appetite or overeating and suicidal thoughts are the symptoms most strongly correlated with MDD-ASCVD. In contrast, poor appetite or overeating is among the least genetically correlated symptoms to MDD (Extended Data Fig. 4c).

Finally, we estimated genetic correlation of attention deficit and hyperactivity disorder (ADHD), anxiety disorders, post-traumatic stress disorder, bipolar disorder, and schizophrenia, with MDD, MDD-ASCVD, and ASCVD. We found that ADHD, anxiety disorder, and PTSD were genetically correlated with ASCVD (Extended Data Fig. 4d). In addition, PTSD and ADHD showed similar genetic correlations for MDD and MDD-ASCVD, which might suggest that the variants that are common to MDD and ASCVD explain most of the genetic correlation between MDD and these disorders.

### Polygenic overlap between MDD and risk factors

Next, we aimed to identify risk factors that contribute to the genetic and phenotypic association between MDD and CVD. First, we assessed the genome-wide genetic correlations between MDD and risk factors. Confirming previous findings, we observed strong to moderate genetic correlations of MDD with psychosocial/lifestyle factors, such as loneliness (*r_g_*=0.68, SE=0.02), childhood maltreatment (*r_g_*=0.55, SE=0.02), and exercise (*r_g_*=-0.33, SE=0.02) (Fig. 5, Extended Data Fig. 6). Among metabolic factors, MDD showed the strongest genetic correlation with type II diabetes (*r_g_*=0.19, SE=0.018), followed by levels of high-density lipoprotein cholesterol (*r_g_*=-0.14, SE=0.01) and triglycerides (*r_g_*=0.18, SE=0.02). MDD showed weak but statistically significant genetic correlations with other metabolic factors (i.e., BMI, non-high-density lipoprotein, low-density lipoprotein, and total cholesterol) (*r_g_*<0.15). We observed significant genetic correlations for MDD with the inflammatory markers IL-6 (*r_g_*=0.22, SE=0.06) and C-reactive protein (*r_g_*=0.15, SE=0.02). We did not observe genetic correlation of MDD with blood pressure traits. As a comparison, for CVDs, the largest genetic correlations were found between heart failure and BMI (*r_g_*=0.55, SE=0.03) and type II diabetes (*r_g_*=0.49, SE=0.03) (Extended Data Fig. 5, Supplementary Table 2). Results for MDD symptoms largely followed the pattern of MDD diagnosis, although changed appetite showed stronger genetic correlations with metabolic factors than did MDD diagnosis (Extended Data Fig. 6, Supplementary Table 3).

Causal variant and local genetic correlation analysis revealed several distinct patterns of polygenic overlap between MDD and risk factors. We found that psychosocial/lifestyle factors, childhood maltreatment, and BMI showed similar levels of polygenicity to MDD (Extended Data Fig. 2b, Supplementary Tables 5 and 7). In addition, they exhibited a large degree of shared variants and many local genetic correlations with MDD (Fig. 4a, Extended Data Fig. 7, Supplementary Tables 4 and 6). Out of these factors, loneliness and childhood maltreatment also showed high levels of effect direction concordance (93% and 73% of shared causal variants and 85% and 75% of significant local genetic correlations for loneliness and childhood maltreatment were in the same direction; Fig. 4b, correlation coefficients are shown in Supplementary Tables 4 and 6). Combined with high polygenicity, such concordance translates to large genome-wide genetic correlations. In contrast, educational attainment, smoking, exercise, physical activity, BMI, and sleep duration had similar levels of polygenicity and a large degree of polygenic overlap with MDD, but low effect direction concordance, suggesting that genome-wide genetic correlations underestimate the polygenic overlap with MDD for these traits.

**Figure 4.**
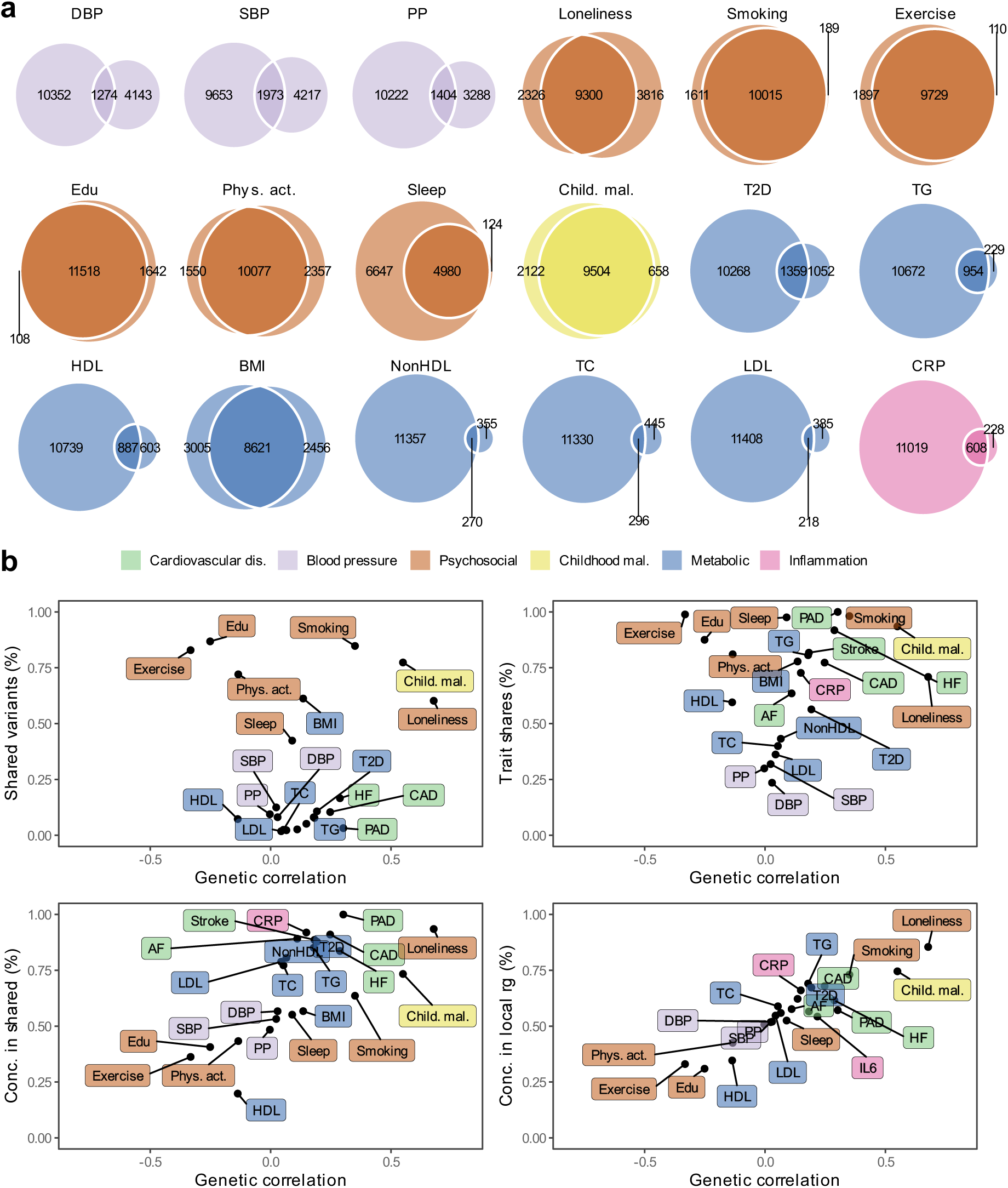
Local and causal-variant level genetic correlations between MDD and risk factors. The legend is shared among all panels. (**a)** Venn diagrams based on MiXeR results showing the number of causal variants that are unique to MDD (left circle), the risk factor (non-overlapping part of right circle) or shared between MDD and the risk factor (overlapping part of circles). (**b)** Genome-wide genetic correlation estimated by LDSC (rg, x-axis) against the percentage of MDD causal variants that are shared with the risk factor as estimated by MiXeR (top left), the percentage of risk factor causal variants that are shared with MDD (top right), the percentage of risk factor causal variants that are shared with MDD that have concordant effect directions (bottom left). The bottom right plot shows the percentage of local genetic correlations from LAVA that have concordant effect directions on the y-axis. Cardiovascular traits are also shown for comparison. **a, b** Standard errors for MiXeR, LAVA, and LDSC results are reported in Supplementary Tables 2-6. Sample sizes for GWAS summary statistics are reported in Supplementary Table 1. Note that IL6 was excluded from MiXeR results because it failed performance checks (see methods). Psychosocial=Psychosocial/Lifestyle; PAD=Peripheral Artery Disease, CAD=Coronary Artery Disease, AF=Atrial Fibrillation, HF=Heart Failure, DBP=Diastolic Blood Pressure; SBP=Systolic Blood Pressure; PP=Pulse Pressure; Edu=Educational attainment; Phys. Act.=Physical activity; Child. Mal.=Childhood Maltreatment; T2D=Type II Diabetes; TG=Triglycerides; HDL=High-Density Lipoprotein; NonHDL=Non-High-Density Lipoprotein; TC=Total Cholesterol; LDL=Low-Density Lipoprotein; IL6=Interleukin-6; CRP=C-Reactive Protein;

Genetic factors for blood pressure traits showed unique patterns of polygenic overlap in that they were polygenic (>5,000 causal variants; Extended Data Fig. 2b) but did not overlap strongly with genetic risk factors for MDD (<0.30 of risk variants overlapping; *Fig.* 4a). Moreover, risk variants that did overlap showed low degree of effect direction concordance (48%-57% of shared variants in the same direction; Fig. 4b). We observed 97 significant local genetic correlations for the three blood pressure traits, 60% of which were positive. These findings suggest that MDD and blood pressure share variants that exhibit both positive and negative correlations, which are cancelled out in the genome-wide estimate.

Type II diabetes, lipid traits, and C-reactive protein showed low polygenicity (<2,500 causal risk variants). Total cholesterol, non-high-density lipoprotein, and low-density lipoprotein did not share the majority of their risk variants with MDD, and shared variants showed low degrees of effect direction concordance. However, like the CVDs, type II diabetes, triglyceride levels, and C-reactive protein levels shared most of their risk variants with MDD, and these variants showed high degrees of concordance (>85% of shared variants in the same direction). High-density lipoprotein shared most of its risk variants with MDD, in consistently opposite directions.

### Risk factors explain part of the genetic correlation between MDD and CVD

To assess the degree to which risk factors explain the genetic overlap between MDD and CVD, we estimated genetic correlations adjusted for risk factors (individually or as a group) using Genomic SEM (Fig. 5a; results per individual risk factor are shown in Extended Data Fig. 8 and Supplementary Table Table 16). The largest reduction in point estimate was observed for the genetic correlation between MDD and peripheral artery disease after adjustment for the group of psychosocial/lifestyle factors, indicating that these risk factors explain much of the genetic correlation between MDD and peripheral artery disease. Similarly, the genetic correlation between MDD and coronary artery disease, as well as the latent CVD factor, were attenuated after adjusting for psychosocial/lifestyle factors. The reduction was mainly driven by loneliness (Extended Data Fig. 8). Genetic correlations of MDD with peripheral artery disease and stroke were no longer significant after adjusting for psychosocial/lifestyle factors. We also observed some attenuation in the genetic correlation between MDD and CVD after adjusting for childhood maltreatment, metabolic factors, or inflammatory markers, although confidence intervals overlapped.

**Figure 5.**
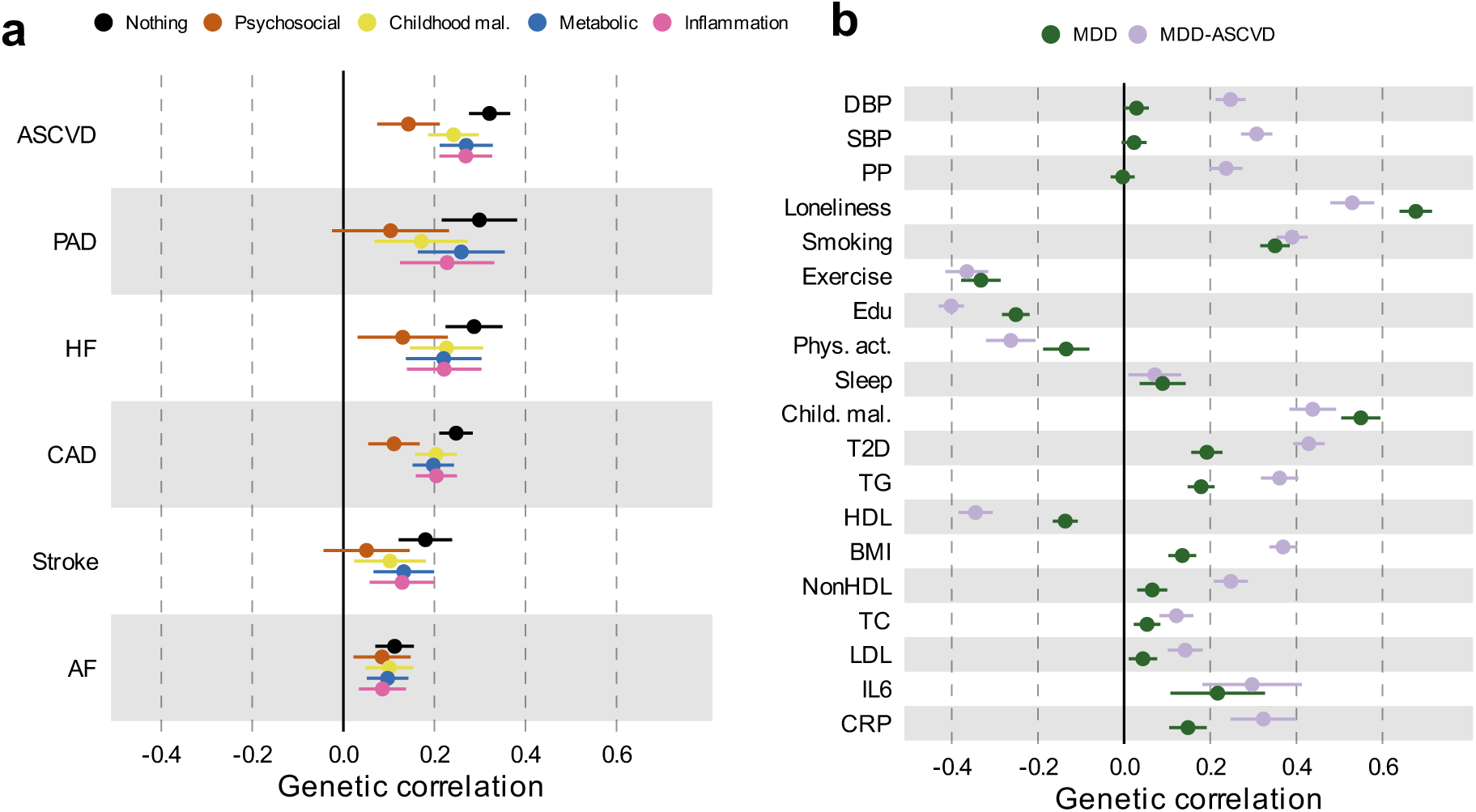
Genome-wide correlations between MDD and risk factors. (**a)** Genetic correlation between MDD and CVD before and after adjustment for groups of risk factors (color coded). (**b)** Comparison of genetic correlation between MDD (dark green) and the latent CVD-ASMDD factor (lilac) and individual risk factors. **a, b** Points and error bars represent correlations and 95% CIs. Sample sizes for GWAS summary statistics are reported in Supplementary Table 1. Psychosocial=Psychosocial/Lifestyle; MDD=Major Depressive Disorder; AF=Atrial Fibrillation; CAD=Coronary Artery Disease; HF=Heart Failure; PAD=Peripheral Artery Disease; DBP=Diastolic Blood Pressure; SBP=Systolic Blood Pressure; PP=Pulse Pressure; Edu=Educational attainment; Phys. Act.=Physical activity; Child. Mal.=Childhood Maltreatment; T2D=Type II Diabetes; TG=Triglycerides; HDL=High-Density Lipoprotein; NonHDL=Non-High-Density Lipoprotein; TC=Total Cholesterol; LDL=Low-Density Lipoprotein; IL6=Interleukin-6; CRP=C-Reactive Protein

Next, we specified the risk factors as mediators instead of covariates in the Genomic SEM model and compared the path estimates from MDD to CVD. Observing attenuation in the association between MDD and CVD when a risk factor is modelled as a mediator rather than a covariate supports the interpretation that the risk factor mediates some of the link between MDD and CVD. For all psychosocial/lifestyle factors together, the inflammation traits, and for type II diabetes, the 95% confidence intervals did not overlap between the mediator and covariate models, suggesting that these risk factors are mediating part of the link between MDD and CVD (Supplementary Table 17).

Finally, we estimated genetic correlations between the risk factors and the latent MDD-ASCVD factor (Fig. 5b) and found that the latent MDD-ASCVD factor was substantially more genetically correlated with blood pressure traits, C-reactive protein levels, and metabolic factors than MDD only, suggesting that these factors characterize the genetic liability to MDD-ASCVD rather than to MDD alone.

### Causal pathways linking MDD and CVD

We investigated putatively causal relationships between MDD and CVD using two-sample MR. Results from the inverse variance-weighted (IVW) estimator are presented in Fig. 6a-c (full results including sensitivity analyses Supplementary Table 18). Instruments were Steiger filtered, i.e., SNPs explaining significantly more variance in the outcome than the exposure were excluded. The results provide support for a causal effect of MDD liability on all CVDs, with the strongest effects observed for coronary and peripheral artery disease. There was no statistically significant pleiotropy for the CVD outcomes, and the results were consistent across weighted median, mode, and Egger sensitivity analyses, providing support for the IVW estimates. Concerning risk factors, we observed no support for a causal effect of MDD liability on levels of blood pressure, physical activity, sleep duration, or IL6. However, increased liability to MDD was associated with increased risk of loneliness, smoking, type II diabetes, and levels of C-reactive protein (Fig. 6a). The results were again consistent across sensitivity analyses. There was statistically significant pleiotropy between MDD and triglycerides, rendering this effect uninterpretable. The average instrument strength of MDD was F=33. Heterogeneity across instruments was statistically significant for most outcomes, suggesting variable effects (Supplementary Table 18).

**Figure 6.**
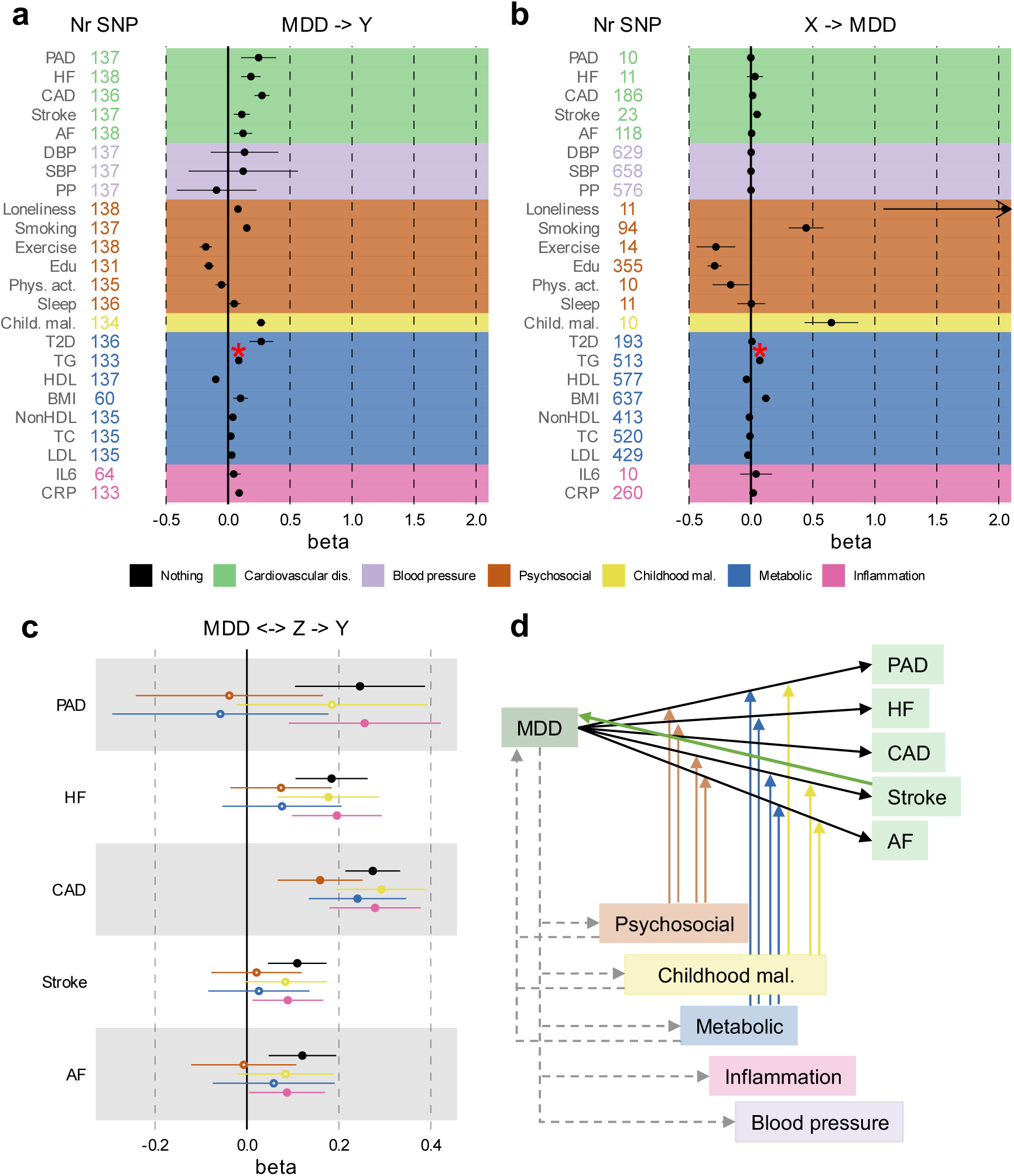
Results from univariable and multivariable MR (IVW estimates). (**a)** Effect of liability to MDD (exposure) on CVD and risk factors (outcomes). (**b)** Effects in the opposite direction with MDD as outcome and CVD and risk factors as exposures. (**c)** Effects of MDD on CVD while adjusting for groups of risk factors in multivariable MR. (**d)** Schematic overview of levels of evidence for causal effects, with solid lines indicating convincing evidence (consistent across sensitivity analyses) for such effects, and dashed lines indicating evidence for some of the relationships tested within the trait categories. The arrows from the risk factors to the association between MDD and the CVDs indicate that the combined risk factors attenuated the association so that it was no longer statistically significant. **a-d** Points and error bars represent regression coefficients and 95% CIs. Sample sizes for GWAS summary statistics are reported in Supplementary Table 1. The term beta refers to the log odds ratio. * Indicates that the observed statistically significant association suffered from pleiotropy; possible causal effect should not be interpreted. Psychosocial=Psychosocial/Lifestyle; MDD=Major Depressive Disorder; MDD-ASCVD=common factor for CVD and MDD and ASCVD; PAD=Peripheral Artery Disease; HF=Heart Failure; STR=Stroke; CAD=Coronary Artery Disease; AF=Atrial Fibrillation; DBP=Diastolic Blood Pressure; SBP=Systolic Blood Pressure; PP=Pulse Pressure; Edu=Educational attainment; Phys. Act.=Physical activity; Child. Mal.= Childhood Maltreatment; T2D=Type II Diabetes; TG=Triglycerides; HDL=High-Density Lipoprotein; NonHDL=Non-High-Density Lipoprotein; TC=Total Cholesterol; LDL=Low-Density Lipoprotein; IL6=Interleukin-6; CRP=C-Reactive Protein; MR=Mendelian randomization IVW=Inverse Variance Weighted.

We also investigated the potential causal effect in the other direction with genetic instruments to CVDs and risk factors as exposures and MDD as outcome (Fig. 6b). We provide evidence for a statistically significant causal effect of stroke, loneliness, smoking, exercise, educational attainment, childhood maltreatment, high-density lipoprotein, and BMI on MDD risk (Fig. 6b), which was robust across the sensitivity analyses. No robust effects were observed for other CVDs, blood pressure, other metabolic traits, or inflammatory markers.

When using genetic instruments for the latent MDD-ASCVD factor to predict the risk factors, statistically significant effects were observed for pulse pressure and type II diabetes (Extended Data Fig. 9c). There was statistically significant pleiotropy only for systolic blood pressure. There were small effects on BMI, childhood maltreatment, educational attainment, high-density lipoprotein, exercise, loneliness, and smoking, but these were less robust across sensitivity analyses (Supplementary Table 18). For most outcomes, there was statistically significant instrument heterogeneity.

We extended univariable results with multivariable MR to assess if there was support for causal effects of MDD on CVD explained by the risk factors. We included only risk factors that were statistically significantly predicted by MDD in the univariable MR analysis. Fig. 6c shows that, as in the Genomic SEM analyses, the effect of MDD on CVD was attenuated after adjusting for groups of risk factors, although confidence intervals were wide. Results for individual risk factors are shown in Extended Data Fig. 10 and Supplementary Table 19. The effect of MDD liability on peripheral artery disease, heart failure, stroke, and atrial fibrillation risk was no longer statistically significant after adjusting for the groups of metabolic or psychosocial/lifestyle factors, although the confidence intervals overlapped with those from the unadjusted estimates. Likewise, the effect of MDD on atrial fibrillation was no longer statistically significant after adjusting for childhood maltreatment. In Fig. 6d we show a schematic overview of results from all MR analyses.

To investigate possible bias due to sample overlap, we repeated the main IVW analyses using MDD summary statistics based on GWAS excluding the UKB sample (the main source of overlap). We observed small differences in the estimates, but the interpretation remained the same for all results (Extended Data Fig. 9a-b, Supplementary Table 20). Furthermore, we repeated the analyses using Latent Heritable Confounder (LHC) MR, which is robust to sample overlap^31^. The results pattern was similar, although the point estimates were slightly attenuated for CVD risk (e.g., no longer statistically significant for heart failure and atrial fibrillation) and became stronger for most other traits (Extended Data Fig. 9c). We conclude that although sample overlap impacted the point estimates, the interpretation of results remained similar.

## Discussion

Here, we showed that genetic risk factors for CVD overlap strongly with MDD. We modeled the shared genetic liability between MDD and ASCVD as a latent factor and showed that, distinct from MDD alone, it is associated with gene expression specific to thalamic and vascular cell-types in the brain and is genetically correlated with immunometabolic factors and blood pressure. Further, we showed that the association between MDD and CVD is partly explained by modifiable risk factors and provide evidence that it is likely causal in nature.

In line with previous results^32–35^ we found weak to moderate genome-wide genetic correlations between MDD and CVDs. Analysis on the level of shared risk variants showed that MDD was substantially more polygenic than the CVD traits and that most risk variants for CVD were in fact shared with MDD and had concordant effect directions. In addition, we found many positive local genetic correlations between MDD and CVD, especially for coronary artery disease although that might reflect the fact that the coronary artery disease GWAS have substantially larger effect sample size than the GWASs of the other CVDs. Interestingly, we found that the *HLA* region was a hotspot for local genetic correlation between MDD and CVDs. These findings suggest that genetic overlap between MDD and CVD is underestimated in genome-wide correlation analyses.

We modelled the shared liability between MDD and ASCVD as a latent factor and performed a GWAS on the factor. Atrial fibrillation deteriorated the fit of the latent factor and it was therefore excluded. This was in line with results that showed that atrial fibrillation was substantially less genetically correlated with the ASCVDs than they were with each other. Combined, these results suggest that atrial fibrillation and ASCVDs have partly distinct sources of genetic variation. We identified many loci for MDD-ASCVD, some of which were uniquely associated with the shared liability and not with the constituent traits. We found that heritability for the MDD-ASCVD latent factor was enriched for genes specifically expressed in vascular braincells, deep layer corticothalamic 6b (projecting to the thalamus), and midbrain-derived inhibitory neurons (predominantly located in the thalamus). This cell-type enrichment signature was not found for MDD or ASCVD alone, suggesting that a distinct mechanism involving thalamic circuits underlies the shared liability to MDD-ASCVD. Altered thalamic function has indeed been implicated previously in CVD^36–38^ and MDD^39,40^, and white matter integrity in thalamic radiations show associations with aortic area and myocardial wall thickness, suggesting that it has a role in the ‘heart-brain’ connection^41^.

MDD showed a high degree of genetic overlap with risk factors. We showed that MDD was substantially more polygenic than blood pressure traits, lipid traits, and C-reactive protein. In contrast, psychosocial/lifestyle traits were equally polygenic to MDD and showed a large degree of overlap with MDD. Interestingly, the local and variant-level analysis indicated that blood pressure traits shared a substantial proportion of their risk variants with MDD (in line with a previous report^42^), which was masked at the genome-wide level due to their opposing effect directions. Similarly, BMI and lipid traits showed discordant effect directions to MDD in overlapping risk variants as well as opposite directions in local genetic correlations. Finally, C-reactive protein shared almost three quarters of its risk variants with MDD, most of which were in the positive direction, indicating a genetic relationship between C-reactive protein and MDD that was masked by the large polygenicity of MDD. Overall, these findings refine our understanding of the polygenic overlap between MDD and risk factors shared between MDD and CVD, and indicate that it is stronger and more complex than has previously been reported.

We estimated genome-wide genetic correlations between MDD and CVD adjusting for risk factors and found that psychosocial/lifestyle factors explain a substantial part of the genetic correlation between MDD and CVD and highlight loneliness as an important factor in the relationship between MDD and CVD.

We found that, compared to MDD, the latent MDD-ASCVD factor was characterized by genetic correlations with immunometabolic factors and blood pressure, suggesting that the shared genetic liability to MDD and ASCVD is associated with an immunometabolic subtype of depression. The existence of an immunometabolic subtype of depression has been proposed previously, based on a long line of evidence of oxidative stress and neuroendocrine and inflammatory dysregulation in MDD, that are preferentially associated with atypical symptoms of MDD (e.g., weight gain and oversleeping)^43^. Indeed, we find that poor appetite or overeating is the MDD symptom with the consistently greatest genetic correlations to the CVDs, and it is among the most genetically correlated with MDD-ASCVD, while it is among the least correlated symptoms to MDD, although confidence intervals were too wide to be conclusive. We did not find large genetic correlations between sleep duration measured using an accelerometer over one week and the MDD-ASCVD factor. However, statistically significant variants for the MDD-ASCVD factor were strongly enriched in statistically significant variants for self-reported short sleep duration (<6 hours per night)^44^ suggesting that short sleep duration is more related to MDD-ASCVD comorbidity than overall sleep duration.

Mental disorders that are highly comorbid with MDD, such as psychotic disorders, anxiety disorders and PTSD, have also been shown to be associated with CVD^45^. We find that ADHD, anxiety disorders and PTSD show genetic correlations with ASCVD and MDD-ASCVD. For ADHD and PTSD, the genetic correlation was similar between MDD and MDD-ASCVD. Future work should estimate latent factors representing shared and distinct sources of genetic covariance between MDD, ADHD, anxiety disorders, and PTSD and investigate how those factors relate to ASCVD.

We found robust support for the likelihood of causal effects of MDD on CVD. Previous two-sample MR studies have observed associations between genetic liability to MDD and risk of coronary artery disease but results for risk of heart failure have been inconsistent^32,33,46,47^. Another study found an effect of genetic liability to MDD on risk of stroke^48^. Using more recent GWAS data, we confirmed an effect of genetic liability to MDD on coronary artery disease and stroke and found robust associations for heart failure and peripheral artery disease.

Except for stroke, we found limited evidence for a causal effect of CVDs on MDD, which is contrary to literature suggesting that such effects exist^24^. The MDD sample is mainly based on large volunteer-based studies that might select against individuals with CVD. Indeed, participants in the UKB study are healthier than the general population^49^. In addition, interpretation of the MR estimate in this case is complicated by the fact that CVD is a time-varying exposure with late age-of-onset^50^. However, we do find that genetic instruments capture the well-established association between stroke and subsequent MDD^51^. Therefore, the findings in our study offer some indication that the association between CVDs and subsequent MDD might have been overestimated in previous studies, possibly due to reverse causation, surveillance bias, or unmeasured confounding.

We observed effects of genetic liability to MDD on most of the risk factors. We did not observe associations between genetic liability to MDD and blood pressure traits, although the presence of correlated and anti-correlated genetic components complicates interpretation. Indeed, using genetic instruments for the latent MDD-ASCVD factor, we did observe strong associations for pulse pressure. Previous MR studies of the association of C-reactive protein and IL-6 levels with MDD risk have also shown inconsistent results^52,53^. We did not find an effect of genetic instruments for inflammatory markers on MDD. However, we did find associations between genetic liability to MDD and increased C-reactive protein levels, lipid levels, and type II diabetes, offering evidence that MDD might lead to long-term dysregulated immunometabolic pathways^43^. Likewise, in line with previous evidence^54^, we provided support for a causal effect of liability to MDD on smoking, which, in turn, can lead to inflammation. These results are in line with results based on genetic covariance estimated from all genetic variants that tentatively supported a mediating role for C-reactive protein and IL6 levels in the association between MDD and ASCVD.

Analyzing risk factors explaining the effect of genetic liability to MDD on CVDs, we observed that only the association between genetic liability to MDD and coronary artery disease remained statistically significant after adjusting for psychosocial/lifestyle or metabolic covariates in a multivariable MR analysis. In addition, we found that smoking status attenuates the association between genetic liability to MDD and peripheral artery disease, for which smoking is a particularly strong risk factor^55^. Interestingly, we find that loneliness is an equally important factor explaining the relationship between MDD and peripheral artery disease, as well as heart failure, which is in line with the results from the adjusted genome-wide genetic correlation analysis discussed above. This emphasizes the need for interventions and preventive policies for reducing loneliness in the population, which has a further increased prevalence during the COVID-19 pandemic and has been described as a pandemic itself^56,57^.

For most CVDs, no risk factor group could fully explain the genetic association between MDD and the CVD. This suggests that there are other mechanisms at work as well, which are not captured by the genetic data used in the study. For instance, GWAS measure lifetime genetic risk (up to the point of the maximum age in the sample) and cannot capture dynamic processes of cumulative and interactive risk. Relatedly, the statistical genetic tools we employed cannot formally distinguish between mediation and covariation pathways. Future studies should triangulate our findings using longitudinal observational and experimental data. Follow-up studies could also investigate the out-of-sample predictive power and clinical usefulness of polygenic risk scores for the shared liability to MDD-ASCVD, and evaluate their ability to identify individuals at risk for immunometabolic depression, as we could not investigate this due to sample overlap in the present study. Another limitation is that the bivariate MiXeR model was not able accurately estimate the polygenic overlap between MDD and IL6. The likely reason the MiXeR model failed in this case is because the two traits have very different genetic architectures with MDD being highly polygenic and IL6 being the least polygenic of the traits in the study, combined with the low sample size of the IL6 GWAS. In addition, to assess generalizability, these findings should be replicated with data from different ancestry sources. The lack of large genetic datasets from non-European populations is a crucial limitation that is widely acknowledged and yet difficult to circumvent. Observational studies have shown that MDD and CVDs could demonstrate different associations depending on ancestry^58,59^, and more genetic and non-genetic research is needed to understand such differences.

To our knowledge, this is the first study moving beyond bivariate genetic overlap to investigate the latent genetic liability shared between MDD and ASCVD. Our triangulation of genome-wide, local, and variant-level methods provides compelling evidence that MDD and CVD, and their shared risk factors are more strongly overlapping genetically than has previously been reported. Similarly, using genome-wide, variant-level, and genetic instrument methods we show how shared risk factors explain the likely causal association between MDD and CVD. For both MDD and CVDs, we have used updated GWAS summary statistics, with an up to 3-fold increase in sample size compared to previous reports^32–35^.

This study dissected the architecture of polygenic overlap between MDD and CVD, and their shared risk factors to elucidate mechanisms linking these comorbid diseases. Our findings suggest that the shared genetic liability to MDD and ASCVD has a distinct genomic signature compared to MDD or any CVD separately. Moreover, the shared genetic liability shows stronger genetic correlations with immunometabolic risk factors than MDD alone, in line with the idea of an inflammatory^60^ or immunometabolic^43^ subtype of MDD especially associated with atherosclerotic CVDs, highlighting the role of inflammation in MDD-ASCVD comorbidity. Indeed, we found that the HLA region is a hotspot of local genetic correlation between MDD and the CVD traits, that genetic liability to MDD is associated with C-reactive protein levels, and tentative support that inflammatory markers mediate some of the link between MDD and ASCVD. We highlight loneliness and smoking as important targets for intervention to reduce the risk of MDD and CVD, as well as CVD in individuals with MDD. Building on this work, tools can be developed to identify individuals at risk for developing immunometabolic depression (e.g., using blood tests of high-density lipoprotein and C-reactive protein levels) and target them for cholesterol-lowering or anti-inflammatory medical interventions.

## Methods

### Data sources

All data sources were summary statistics from the largest and most recent GWAS to date (see Supplementary Table 1). The MDD symptoms GWAS have not been published, although they have been used in other publications^61^. The symptom GWASs were based on Patient Health Questionnaire (PHQ-9) items measured in the UKB, that captures most, but not all, core symptoms of MDD, and were published online^62^. The disease trait GWASs were mostly based on electronic health record diagnoses. The physical activity and sleep duration traits were measured in the UKB using an accelerometer over a one-week period^63^.

The summary statistics were cleaned using the cleansumstats pipeline (https://github.com/BioPsyk/cleansumstats). SNPs were aligned and harmonized against reference data. Analyses were conducted on the TSD cluster, maintained by the University of Oslo, using singularity containers. Containers are packaged applications with their environmental dependencies that are used to standardize analyses across different sites, ensuring correct software versions and parameters^64^. All GWAS studies were ethically approved and were conducted in compliance with ethical guidelines. Ethics approval for the UK-Biobank study was given by the North West Centre for Research Ethics Committee (11/NW/0382). The work described here was approved by UK-Biobank under application number 22224.

### Genetic overlap

#### Genome-wide genetic correlation

To assess shared liability between MDD, CVD, MDD symptoms, and risk factors, we estimated genetic overlap on a genome-wide, polygenic, and local level. First, for the genome-wide level, we estimated genetic correlation between each of the traits using LD score regression (LDSC^29^). We excluded the human leukocyte antigen (*HLA*) region from the main analysis because its complex LD structure can bias both heritability and genetic correlation results^65^. However, a sensitivity analysis including the *HLA* region showed very similar results to the main analysis (not shown). Note that LDSC performs well in the presence of sample overlap.

#### Local genetic correlation

We used Local Analysis of (co)Variant Association (LAVA^26^) to assess genetic correlation in regions of the genome. We assessed local genetic correlation in 2,495 genomic regions that cover the autosomes and have been defined to minimize LD between the regions, while simultaneously keeping the regions approximately equal in size. These regions are provided with the LAVA software package. We only considered local genetic correlation in loci where both traits showed marginally significant heritability (*P*<0.05). For these loci, we adjusted local genetic correlation *P*-values for multiple testing using the Benjamini-Hochberg method. This adjustment was done separately for all pairs of traits considered. To match the results from LDSC and MiXeR, we excluded the *HLA* region from the main analyses. For local genetic correlation between MDD and the CVD traits, we performed an additional analysis in the *HLA* region.

#### Polygenic overlap

To investigate polygenic overlap beyond genetic correlation, we used MiXeR v1.3^25^ to assess the number of shared and distinct non-zero genetic variants between MDD and another trait required to explain at least 90% of heritability in the two traits, referred to as ‘causal’ variants. Because it assesses overlap regardless of the effect direction of each variant, it gives a more adequate picture of localized correlations that could cancel each other out when estimating the genome-wide genetic correlation. We excluded the *HLA* region, following the software recommendations^25^. A sensitivity analysis of genetic overlap between MDD and the CVD traits including the *HLA* region showed very similar results to the main analysis (not shown). To assess stability of point estimates and estimate their standard deviations, we fitted the MiXeR model 20 different times for 2 million randomly selected SNPs with minor allele frequency of at least 5%. The number of 20 runs follows recommendations published previously^66^. The averaged results were then plotted in Venn diagrams to visualize the polygenic overlap. The MiXeR model was evaluated for each trait using by 1) comparing the AIC of the univariate MiXeR model with the LDSC model, which does not include a parameter to directly estimate polygenicity (Supplementary Table 5); 2) comparing the AIC of the bivariate MiXeR model with the AIC of the model with the least possible amount of polygenic overlap required to explain observed genetic correlation; 3) comparing the AIC of the bivariate MiXeR model with the AIC of the model with maximum amount of polygenic overlap (in such a model, all risk variants of the least polygenic trait are also risk variants of the other trait), and finally 4); evaluating the stability of the point estimates over the 20 runs (Supplementary Table 6). These metrics have been described in detail previously^66^. Except for the univariate test, the MiXeR model failed these checks for IL6, which was therefore excluded from the results. The MiXeR model performed poorly for the IL6 likely because it is a trait with low polygenicity and was measured in a relatively small sample (Extended Data Fig. 2, Supplementary Table 1).

To complement the univariate polygenicity analysis in MiXeR, we estimated polygenicity in a Bayesian model implemented in SBayesS^30^. We used a 15,000 sample MCMC chain with a 5,000-sample burn-in. The SBayesS methods did not converge for peripheral artery disease, possibly due to lower number of cases in the GWAS.

### Shared liability to MDD and ASCVD

To move beyond bivariate association to multivariate overlap, we conducted factor analysis on MDD and the CVDs using Genomic SEM^27^ to assess if genetic latent factors could explain the genetic covariance between the traits. Genomic SEM uses LDSC to estimate the genetic covariance matrix and uses that in a SEM framework to identify multivariate relationships in the data. A Comparative Fit Index CFI>.90, Standardized Root Mean Square Residual SRMR<.03, and standardized factor loadings *β*>0.3 were set as criteria for acceptable model fit. The best model fit was found for a factor with coronary artery disease, peripheral artery disease, heart failure, and stroke as indicators, which we interpret as a genetic factor for ASCVD. To model a genetic factor for shared liability to MDD and ASCVD, we defined a higher-order factor with this ASCVD factor plus MDD as indicators. The standardized loading of the first indicator (coronary artery disease) was set to 1. The residual variance of the ASCVD factor was forced to be 0, so that all variance was forced into the MDD-ASCVD factor. For comparison, we also estimated a common factor model for ASCVD without MDD, which is visualized in Extended Data Fig. 3a.

Subsequently, we conducted a summary statistics-based GWAS on the MDD-ASCVD second-order latent factor to identify variants associated with this latent liability. We used the package-default diagonally weighted least squares estimator (DWLS). To derive genome-wide significant independent loci we used the plink clumping procedure as implemented in FUMA^67^, with R=0.6 and distance=250 kb, and reference data from 1000 genomes. To assess the heterogeneity of the SNP effects, we also fit an independent pathway model for each SNP, where each indicator was regressed on the SNP directly instead of forcing the effect through the latent factor. We compare the common pathway 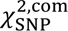 to the independent pathway 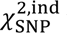 to derive Q_SNP_. For our follow-up analyses, we filtered out all SNPs that had effects that were more consistent with an independent pathway model at *P*_QSNP_ <0.05. This stringent procedure filters out SNPs with heterogenous effects on MDD and the ASCVD factor. This way, variants are excluded that should be considered risk variants for MDD or CVDs separately rather than risk variants for MDD-ASCVD.

Using FUMA, we checked if genome-wide significant SNPs for the MDD-ASCVD factor were enriched in genome-wide significant SNPs for traits in the GWAS-catalog. Then, to define unique loci, we overlaid the independent genomic risk loci (now clumped at R^2^=0.1 and 3000 kb window) for MDD-ASCVD with the risk loci for the constituent traits (MDD, peripheral artery disease, coronary artery disease, heart failure, and stroke). We used the ‘intersect’ function in bedtools to see if the risk regions were overlapping. If they were independent (according to the clump criteria) they were regarded as novel loci. We also identified genes whose regulation is significantly impacted by the top significant SNPs to interpret the biological implications of our findings. For this, we use eQTL estimates from FUMA based on PsychENCODE reference data^68^. Using the same procedures (though without filtering out heterogeneous SNPs) we conducted a GWAS for the the ASCVD genetic factor.

To externally validate the latent MDD-ASCVD GWAS results, we computed PRS using LDpred2 (automatic mode with HapMap3 reference data). We compared the MDD-ASCVD PRS with a PRS based on MDD only, and with a PRS based on latent CVD without MDD. As a target sample we used the UKB. Summary statistics for ASCVD traits excluding the UKB were unavailable, and we chose to leave the UKB in for all traits. Sample overlap is likely to lead to overfitting, resulting in an inflation of explained variance. However, this is less of a concern when comparing PRS among themselves, rather than assessing absolute predictive value. As target phenotypes, we extracted CVD and MDD cases according to healthcare registry ICD codes from data field 41270 (see Supplementary Table 12). We used logistic regression analysis to predict case status from each PRS while controlling for the first 10 principal components for ancestry, sex, and year of birth. Continuous variables were standardized and centered. We estimated Nagelkerke’s R^2^ to capture explained variance in the disease traits.

### Tissue and cell-type analysis

To gain insight into the biological mechanisms underlying the common liability to MDD and ASCVD, we performed a tissue and cell-type analysis using partitioned LDSC. Cell-type identification was based on the top decile of specifically expressed genes (referred to as top decile expression proportion [TDEP] genes). The methodology has been described extensively in previous studies^69–71^.

We identified TDEP genes for brain cell-types from single-nucleus RNA-seq data measured in dissections of three adult human post-mortem brain samples for the Adult Human Brain Atlas^28^, part of the Human Cell Atlas. We used the manually annotated 31 superclusters and 461 clusters provided by the atlas^28^. We considered a curated set of 18,090 protein-coding autosomal genes, excluding those in the *HLA* region (because the method relies on LDSC), with expression in at least one of the 461 cell-clusters.

To establish TDEP genes for 16 human tissues, we utilized bulk RNA-seq data from GTEx v8^72^. In line with previous research, we removed tissues with <100 donors and non-natural tissues (e.g., cell lines) as well as testis tissues (expression outlier)^69^. Prior to analysis in partitioned LDSC, we expanded the boundaries of TDEP genes by 100kb to include possible enhancers or promoters.

We tested the associations between GWAS traits and tissue/cell-types by investigating heritability enrichment within the TDEP genes for each tissue/cell-type. To ensure that annotation enrichment could not be better explained by other overlapping annotations, we adjusted for enrichment in 53 previously defined baseline LDSC annotations (LDSC v1.0.1) of coding, UTR, promoter, intronic, various enhancers, histone marks and other epigenetic marks, genomic regions^71^.

### Pathways of association between MDD and CVD

#### Mediation in Genomic SEM

We employed several different techniques to assess whether the association between MDD and CVD could be explained by shared risk factors. First, we estimated the genome-wide genetic correlations adjusting for the effects of risk factors using Genomic SEM. To aid interpretation, we added the covariates in groups, controlling for all trait groups separately (psychosocial/lifestyle, childhood maltreatment, metabolic, or inflammation traits; see Supplementary Table 1). We only included traits in the covariate groups that showed significant genome-wide genetic correlation with MDD. Note that we did not adjust for blood pressure traits, since none of the blood pressure traits showed genome-wide significant correlation with MDD. Attenuation of genetic correlation after adjustment was taken to indicate that shared risk factors account for some (or all) of the association between MDD and CVD.

Next, we modeled the risk factors explicitly as mediators in Genomic SEM. If the direct effect is attenuated in the mediation model as compared to the covariate model, we view this as tentative support for the existence of mediation (following procedures suggested by the software developers^73^). Note however that this interpretation relies on untestable assumptions. Finally, we also tested the effects of individual risk factors (both as covariates and mediators) instead of grouping them.

#### Univariable and multivariable Mendelian randomization

To test if the associations between MDD and CVD and risk factors were causal, we used Mendelian Randomization (MR). We assessed the effects of MDD on the CVDs and risk factors, the effect of the CVDs and risk factors on MDD, and the effect of latent MDD-ASCVD on risk factors. MR uses genetic variants as instrumental variables to assess the presence of causal effects of an ‘exposure’ on an ‘outcome’. Core assumptions include that the instrumental variables are robustly associated with exposure and are not associated with the outcome (other than through the effect from exposure) or unmeasured confounders. We used the inverse variance weighted (IVW) estimate in the two-sample MR R-package^74^ as our main estimate. As instrument SNPs, we selected independent GWAS hits at *P<*5×10^-8^, R^2^< 0.001, and distance <5Mb. In the analysis of the effect of genetic instruments of peripheral artery disease, physical activity, childhood maltreatment, and IL6 on MDD risk, we allowed instruments with higher *P*-values to be able to reach a total of 10 instruments (*P<*1e-5). For the MDD-ASCVD exposure, we used SNPs that showed no significantly heterogenous effects in the Genomic SEM model (Q_pval_>.05, see above), in order to limit the possibility of pleiotropic effects of this, by nature, heterogenous instrument.

We performed several sensitivity analyses to test and adjust for violation of MR assumptions. All analyses were Steiger filtered, meaning that all SNPs that explained significantly more variance in the outcome than the exposure were excluded as instruments^75^. Weighted median and mode regression were used to correct for effect size outliers that could represent pleiotropic effects^76^. MR-Egger regression was used to assess pleiotropy (pleiotropy leads to a significant intercept) and correct for it (unless I^2^ indicated NOME violation of the NO Measurement Error [NOME] assumption, in which case we did not report MR-Egger results ^77,78^). Second, to assess the strength of our instruments, we used Cochran’s Q to assess heterogeneity in the SNP effects^79^ and the F-statistic to control for weak instrument bias^80^.

Third, we performed sensitivity analyses to gauge the effect of sample overlap in the GWASs that were used. Although sample overlap has been suggested to not greatly impact MR results when the source GWASs have a large sample size and the overlap is limited^81^, we wanted to ensure sample overlap did not lead to bias in our findings. We assessed the genetic covariance intercepts for all MDD-trait pairs from the LDSC analyses and observed that most were more than 1 SD away from 0, indicating that sample overlap was present (Supplementary Table 2). We repeated the analyses with MDD summary statistics leaving out the UKB sample, which is the sample responsible for most of the overlap and compared the results. Also, we repeated the analyses using Latent Heritable Confounder MR (LHC-MR^31^), which aims to correct for the presence of unmeasured heritable confounders as well as sample overlap.

Subsequently, we test the adjusted pathways using multivariable MR (MVMR) analyses. The difference with the mediation test in Genomic SEM is that we now use instrumental variables that corroborate a causal interpretation. Because MVMR relies on simple regression analysis, it cannot formally test mediation; instead, the causal estimate is adjusted for the risk factor. To support a directional interpretation, we included only risk factors as mediators that were significantly affected by MDD according to the univariable analysis results. In MVMR, only an IVW estimate can be derived. Steiger filtering was performed on the exposure-outcome association. These analyses were not replicated in LHC, which does not accommodate multivariable analyses. For ease of interpretation, we again grouped the mediators and adjusted for all mediators in a group concurrently. We only included a mediator in a group if it was significant in the univariate analysis. Additionally, we performed analyses adjusting for single mediators. We selected instruments for each variable in a model by a clumping step with the same parameters as in the univariate MR case (*P<*5×10^-8^, R^2^< 0.001). We then combined instruments for all variables in a model into a single set of instruments and performed another clumping step with the same parameters. These instruments were then aligned to the same effect allele. We estimated the effect of genetic liability to MDD on CVD traits adjusting for covariates using multivariate MR with the MVMR R-package^82^.

## Data availability

Links to download publicly available published GWAS summary statistics data used as inputs in this study are listed in Supplementary Table 1. Single nucleus RNA sequencing data in the adult human brain can be found at https://github.com/linnarsson-lab/adult-human-brain. Researchers can request access to the UK-Biobank data resources at https://www.ukbiobank.ac.uk/enable-your-research/apply-for-access; data for PRS analysis described in this study were accessed under accession number 22224. Gene expression data from human tissues can be found at https://www.gtexportal.org/home/datasets. Summary statistics for MDDCVD are available upon request.

## Code availability

Code used for processing and analyzing data in this work will be uploaded to https://github.com/jacobbergstedt. Code for singularity containers can be found at https://github.com/comorment.

## Acknowledgments

This work was supported by European Union’s Horizon 2020 Research and Innovation Programme (CoMorMent project; Grant #847776). JAP was supported by the the US National Institutes of Mental Health (R01MH123724). NP was supported by the Marie Skłodowska-Curie Actions Grant 801133 (Scientia fellowship) and RCN 300309. JLT is supported by a European Research Council (ERC) Starting grant (UNRAVEL-CAUSALITY, grant number 101076686) and by Foundation Volksbond Rotterdam. CML is part-funded the NIHR Maudsley Biomedical Research Centre at South London and Maudsley NHS Foundation Trust and King’s College London. AMM is supported by the Wellcome Trust (220857/Z/20/Z) and UKRI (MR/W014386/1). ADB was supported by grants from the Lundbeck Foundation (R102-A9118, R155-2014-1724, and R248-2017-2003), the EU H2020 Program (Grant No. 667302, “CoCA”), NIH/NIMH (1R01MH124851-01). OA was supported by the European Union’s Horizon 2020 research and innovation program (grant agreement No 847776 and 964874), Research Council of Norway (223273, 296030, 326813). PFS gratefully acknowledges support from the Swedish Research Council (Vetenskapsrådet, award D0886501), the US National Institutes of Mental Health (R01s MH124871, MH121545, and MH123724), and the Horizon 2020 Programme of the European Union (grant agreement No 847776). LY was supported by the European Research Council (grant agreement ID 101042183). The authors acknowledge HERMES and CARDIoGRAMplusC4D consortia for contributing data and guidance. The PRS analyses have been conducted using the UK-Biobank Resource under Application Number 22224. They were enabled by resources in project sens2017519 provided by the National Academic Infrastructure for Supercomputing in Sweden (NAISS) at UPPMAX, funded by the Swedish Research Council through grant agreement no. 2022-06725. This work was performed on Services for Sensitive Data, University of Oslo, with resources provided by UNINETT Sigma2, the national infrastructure for high performance computing and data storage in Norway.

## Author contributions

This study was conceived by JB, PFS, YL, and FF. It was supervised by YL and FF. JB, JAP, and ZM were responsible for the main analyses, visualization, and the drafting of the manuscript. AH performed the tissue expression analyses with help and supervision from SY, JHL, and PFS, who also helped with the writing and interpretation of these results. NP was involved in providing and cleaning GWAS summary statistic data. DJAS supervised the Genomic SEM analysis and JLT the MR analyses. OF and AS were involved in providing the containerized software used in this project and supervising the MiXeR analyses. JJM, QS, SH, PT, AB, and TW contributed to interpreting findings. CML, AMM, TDA, and ADB were responsible for the MDD GWAS summary statistics and were involved in the interpretation of findings. SH helped with UKB data access and curation for the PRS analyses. SH, UVA, OA, PFS, YL and FF provided suggestions and expert supervision and contributed to the writing. All authors have read and provided feedback on the manuscript.

---

**Extended Data Fig. 1.**
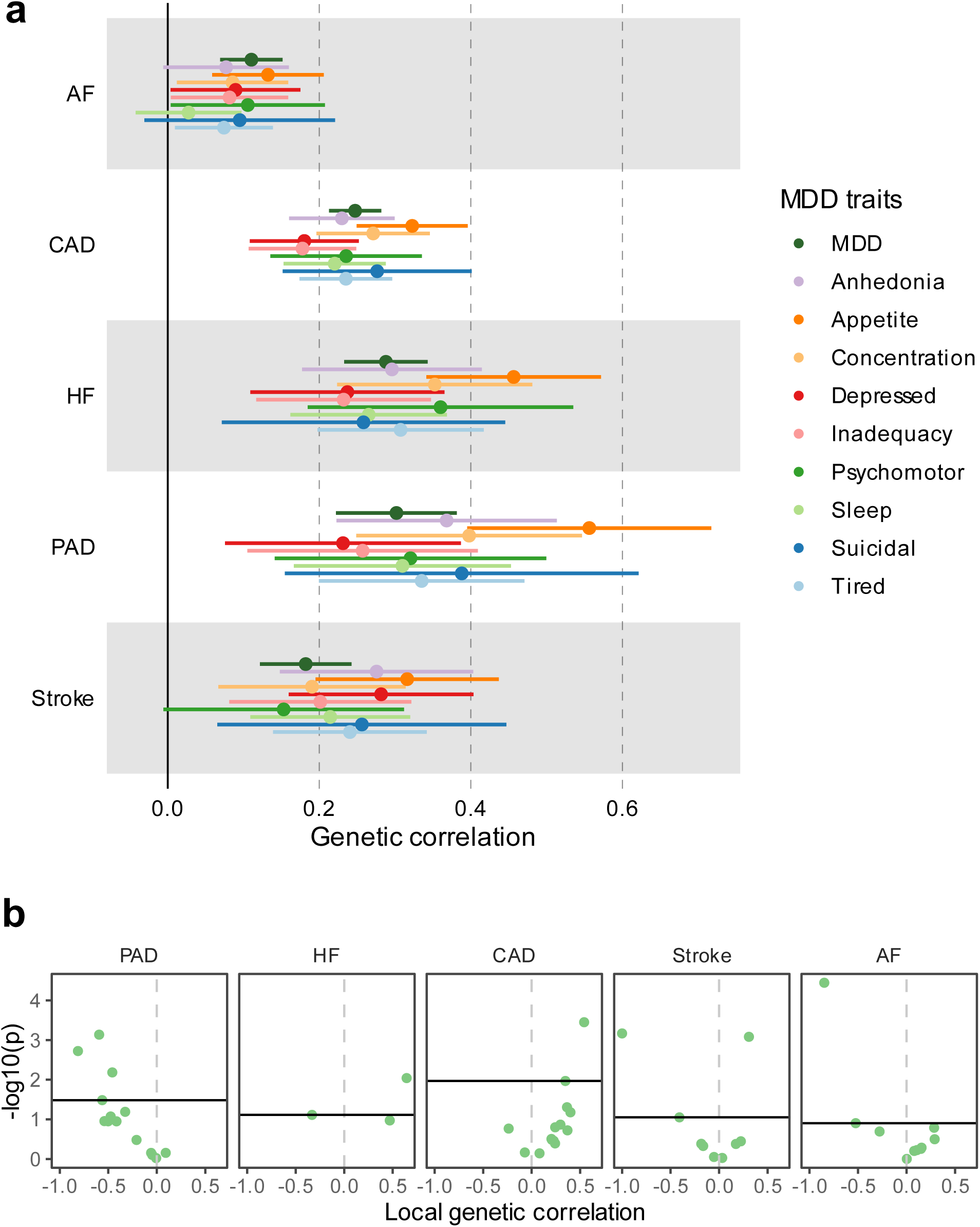
Genetic correlation of MDD and MDD symptoms with CVDs. **(a)** Results are based on LD Score Regression analysis. Points and error bars represent genetic correlation and 95% CIs. Sample sizes for GWAS summary statistics are reported in Supplementary Table 1. **(b)** Local genetic correlation between MDD and CVDs in 16 loci in the HLA region. Only loci with marginally significant local heritability for both traits are shown. Points above the vertical line are significant based on multiple testing adjustment for considered loci performed for each CVD trait separately. MDD=Major Depressive Disorder; AF=Atrial Fibrillation; CAD=Coronary Artery Disease; HF=Heart Failure; PAD=Peripheral Artery Disease

**Extended Data Fig. 2.**
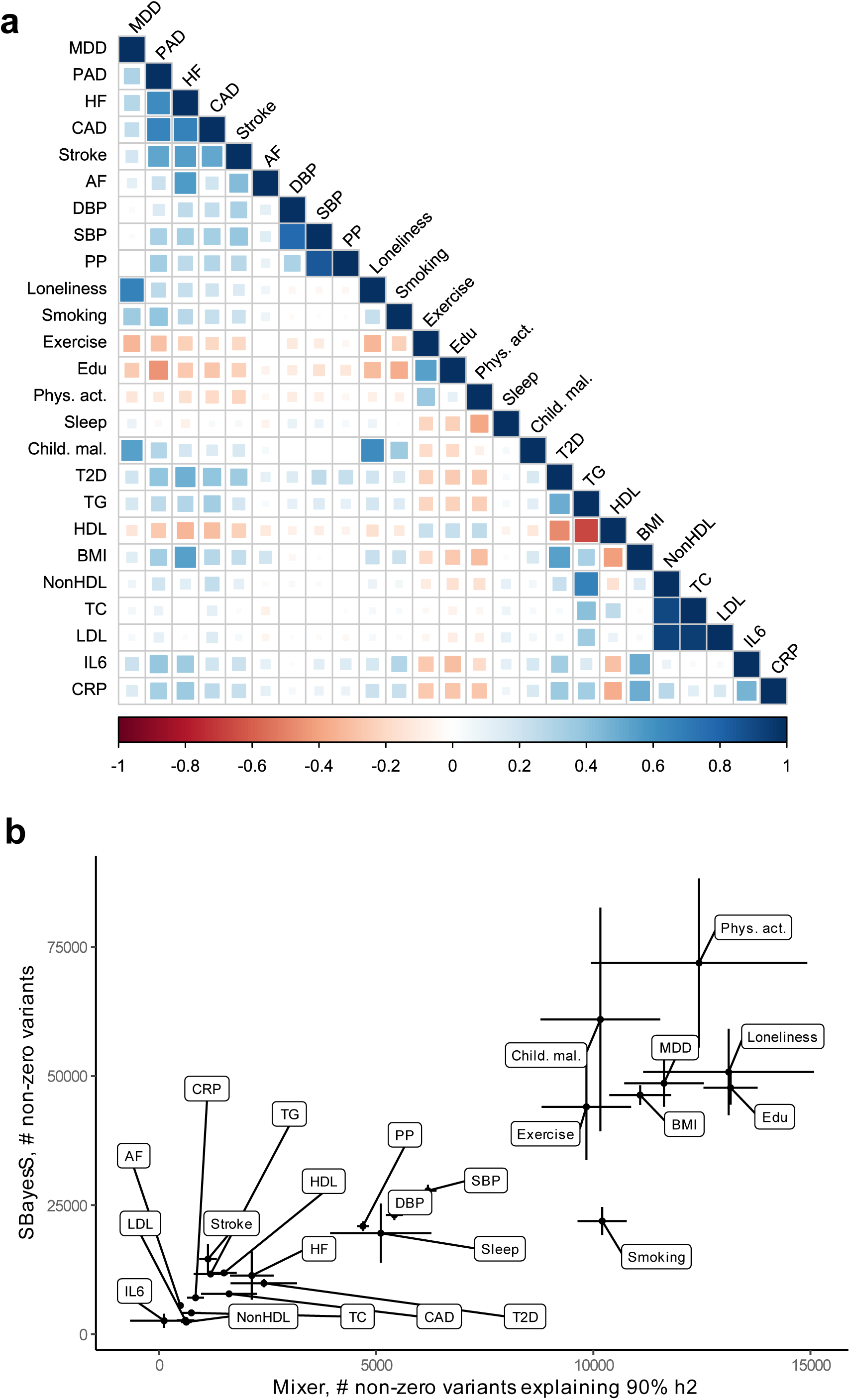
Genetic correlation and polygenicity for MDD, CVD, and risk factors. **(a)** Heatmap of the genetic correlations between MDD, the CVDs, and the risk factors, with the color indicating the effect direction (negative: red, positive: blue) and the size and shade of the square illustrating the size of the correlation. Results are based on LD Score Regression analysis. **(b)** Estimates of number of non-zero variants for from SBayesS for each trait (y-axis) and estimates of non-zero variants required to explain 90% of trait heritability for each trait from MiXeR (x-axis) for MDD, the CVDs, and the risk factors. Note that for PAD polygenicity estimates did not converge for SBayeS, possibly because of few number of cases. MDD=Major Depressive Disorder; AF=Atrial Fibrillation; CAD=Coronary Artery Disease; HF=Heart Failure; PAD=Peripheral Artery Disease; DBP=Diastolic Blood Pressure; SBP=Systolic Blood Pressure; PP=Pulse Pressure; Edu=Educational attainment; Phys. Act.=Physical activity; Child. Mal.=Childhood Maltreatment; T2D=Type II Diabetes; TG=Triglycerides; HDL=High-Density Lipoprotein; NonHDL=Non-High-Density Lipoprotein; TC=Total Cholesterol; LDL=Low-Density Lipoprotein; IL6=Interleukin-6; CRP=C-Reactive Protein

**Extended Data Fig. 3.**
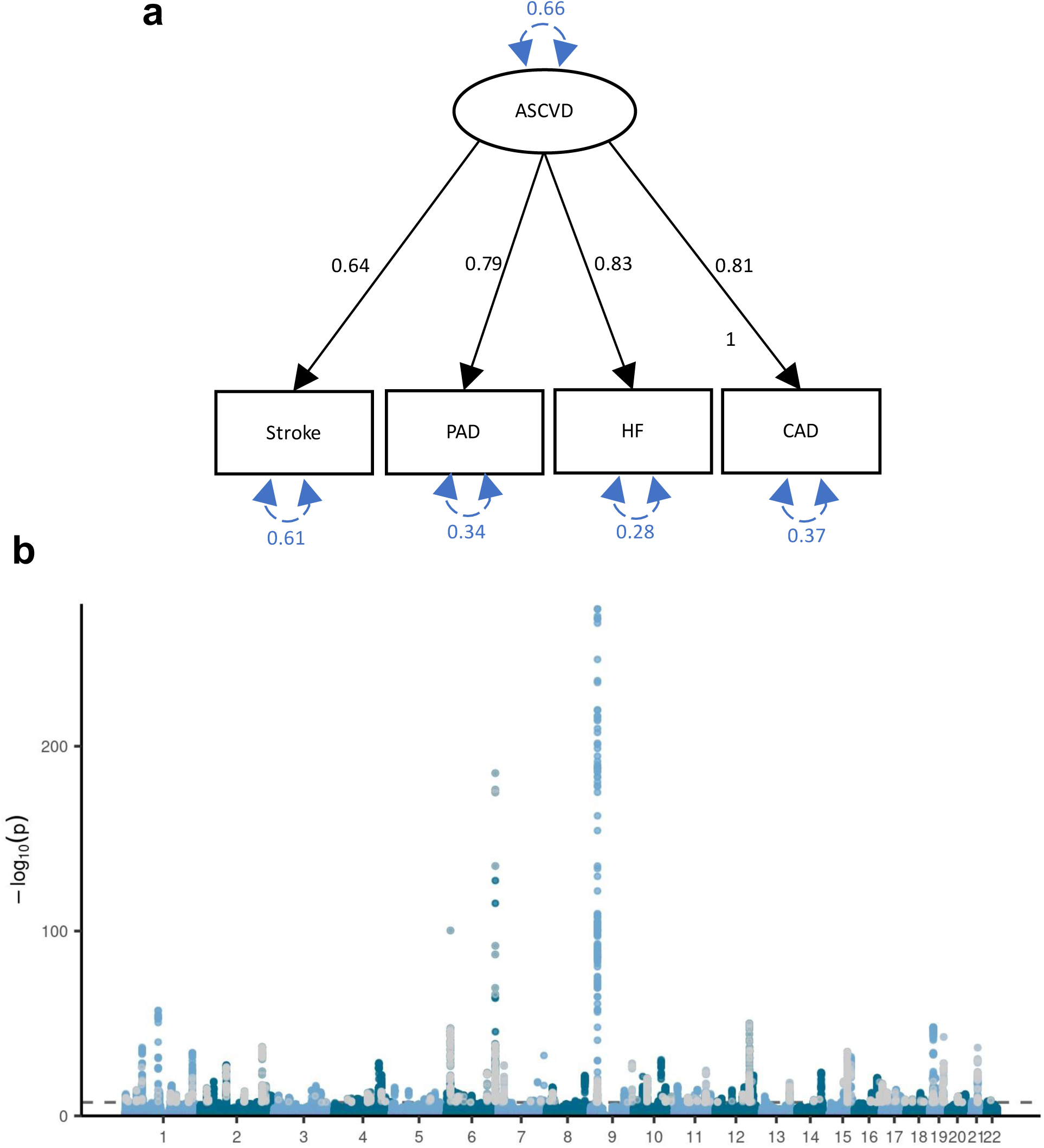
Common latent factor underlying atherosclerotic CVD. **(a)** Latent atherosclerotic CVD (ASCVD) model, defined by stroke, peripheral artery disease (PAD), heart failure (HF), and coronary artery disease (CAD). All ‘observed’ traits are based on GWAS summary statistics. Results are from confirmatory factor analysis in Genomic SEM, and standardized factor loadings are given for each path. Circular dashed arrows give the trait variance. (**b)** Manhattan plot of the GWAS on ASCVD, with each dot representing a SNP with its position on the x-axis and its *P*-value on the y-axis. Genome-wide significant SNPs with a significant heterogeneity Q_SNP_ (with a strong effect on one or some of the indicators that was not well explained through the common latent factor) are displayed in grey. The dashed line indicates the genome-wide significance threshold (*P*<5e-8).

**Extended Data Fig. 4.**
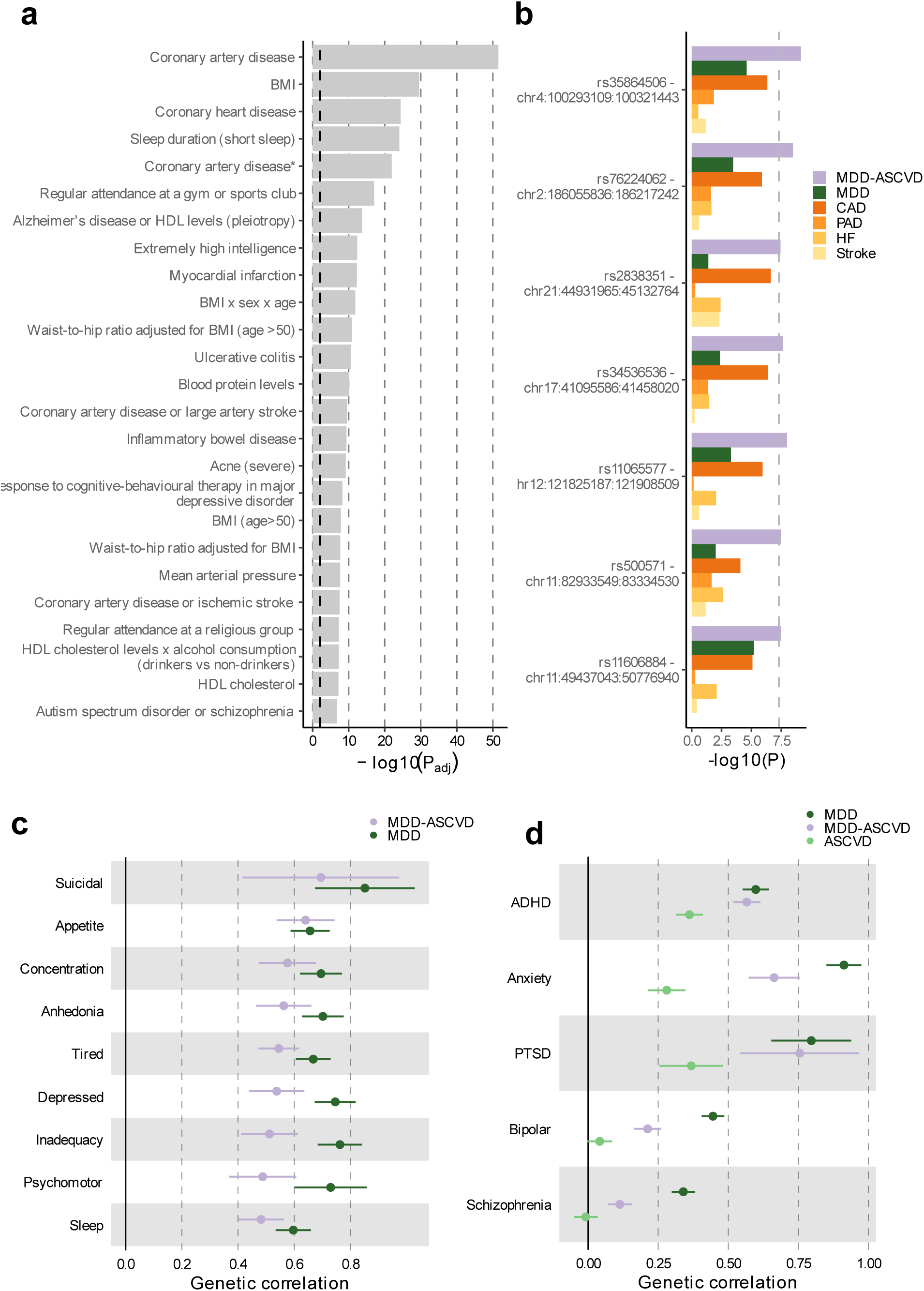
The genomic signature of MDD-ASCVD. **(a)** Enrichment for the MDD-ASCVD GWAS SNPs in genome-wide significant SNPs for traits in the GWAS catalog computed using FUMA. The traits are as reported in the original study. Note that sleep duration here is a dichotomization of self-reported sleep. Full name in GWAS catalog of the trait “Coronary artery disease*” is “Coronary artery disease (myocardial infarction, percutaneous transluminal coronary angioplasty, coronary artery bypass grafting, angina or chromic ischemic heart disease)”. The dashed black line indicates the significance threshold after Benjamini-Hochberg adjustment. Adjustment for multiple testing was conducted over all traits in the GWAS catalog (>3500 traits). **(b)** The five SNPs that were significantly associated with MDD-ASCVD, but not with any of the constituent traits, with their *P*-value in the GWAS of the constituent traits. The dashed line indicates the genome-wide significance threshold (*P*<5e-8). **(c)** Genetic correlation of PHQ-9 MDD symptoms with MDD, and MDD-ASCVD, with bars representing 95% confidence intervals. Results are based on LD Score Regression analysis. **(d)** Genetic correlation of MDD, MDD-ASCVD, and ASCVD with five psychiatric disorders, with bars representing 95% confidence intervals. Results are based on LD Score Regression analysis. ADHD = Attention Deficit and Hyperactivity Disorder; PTSD = Posttraumatic Stress Disorder.

**Extended Data Fig. 5.**
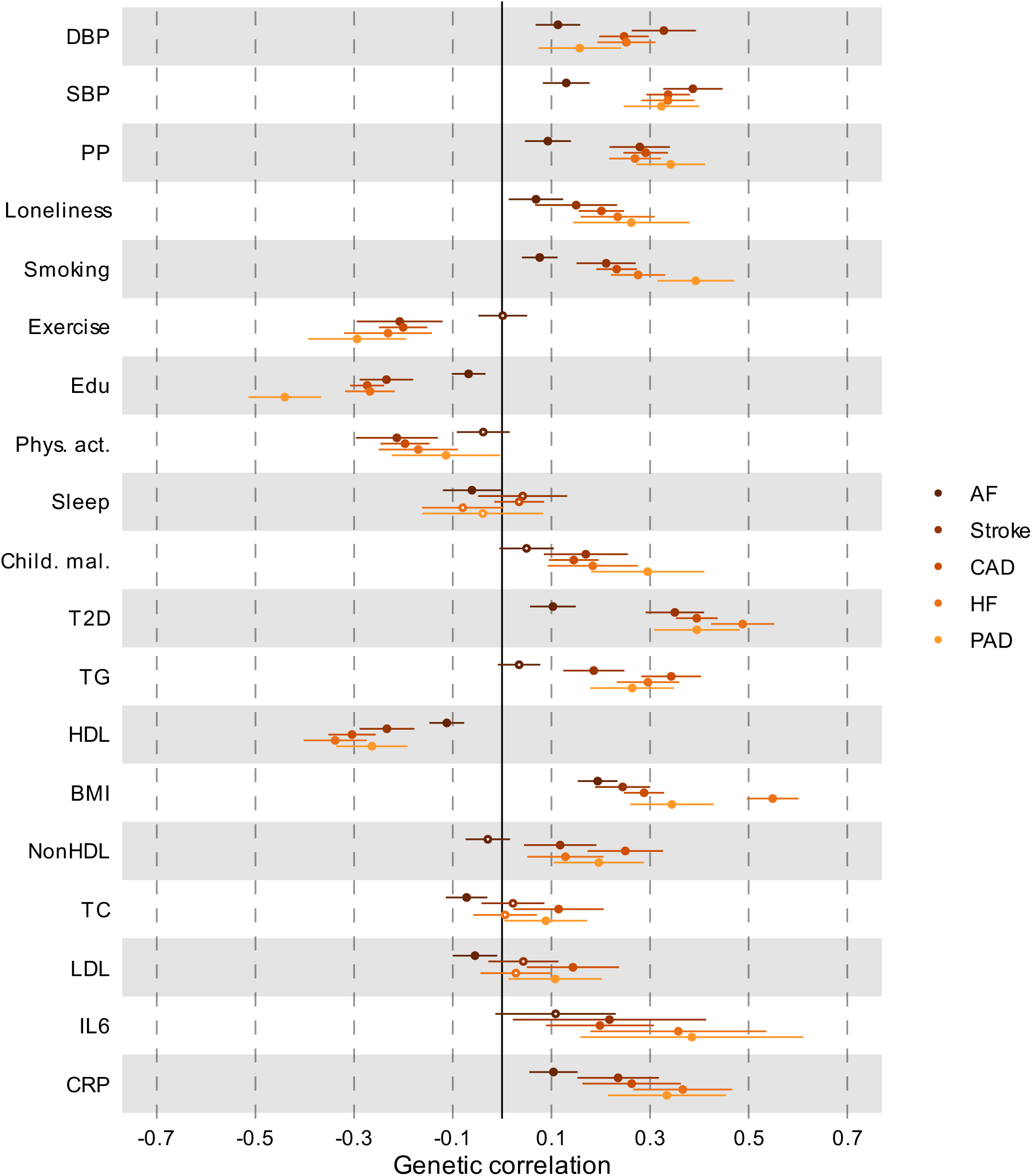
Genetic correlation between CVD and risk factors. Results are based on LD Score Regression analysis. Points and error bars represent correlations and 95% CIs. Sample sizes for GWAS summary statistics are reported in Supplementary Table 1. Open dots indicate a non-significant genetic correlation. AF=Atrial Fibrillation; CAD=Coronary Artery Disease; HF=Heart Failure; PAD=Peripheral Artery Disease; DBP=Diastolic Blood Pressure; SBP=Systolic Blood Pressure; PP=Pulse Pressure; Edu=Educational attainment; Phys. Act.=Physical activity; Child. Mal.=Childhood Maltreatment; T2D=Type II Diabetes; TG=Triglycerides; HDL=High-Density Lipoprotein; NonHDL=Non-High-Density Lipoprotein; TC=Total Cholesterol; LDL=Low-Density Lipoprotein; IL6=Interleukin-6; CRP=C-Reactive Protein

**Extended Data Fig. 6.**
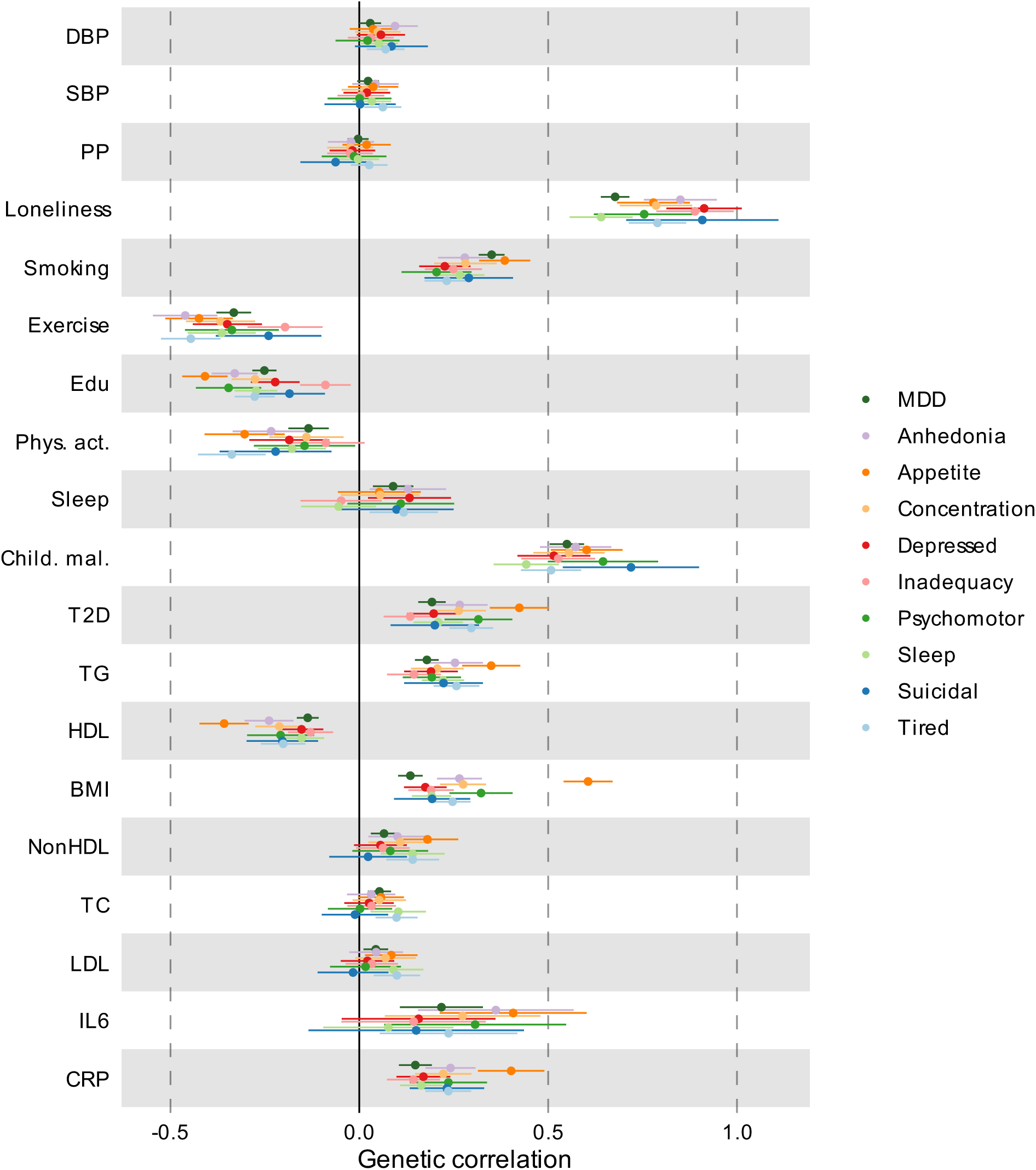
Genetic correlation between MDD traits and risk factors. Results are based on LD Score Regression analysis. Points and error bars represent correlations and 95% CIs. Sample sizes for GWAS summary statistics are reported in Supplementary Table 1. Open dots indicate a non-significant genetic correlation. MDD=Major Depressive Disorder; DBP=Diastolic Blood Pressure; SBP=Systolic Blood Pressure; PP=Pulse Pressure; Edu=Educational attainment; Phys. Act.=Physical activity; Child. Mal.=Childhood Maltreatment; T2D=Type II Diabetes; TG=Triglycerides; HDL=High-Density Lipoprotein; NonHDL=Non-High-Density Lipoprotein; TC=Total Cholesterol; LDL=Low-Density Lipoprotein; IL6=Interleukin-6; CRP=C-Reactive Protein

**Extended Data Fig 7.**
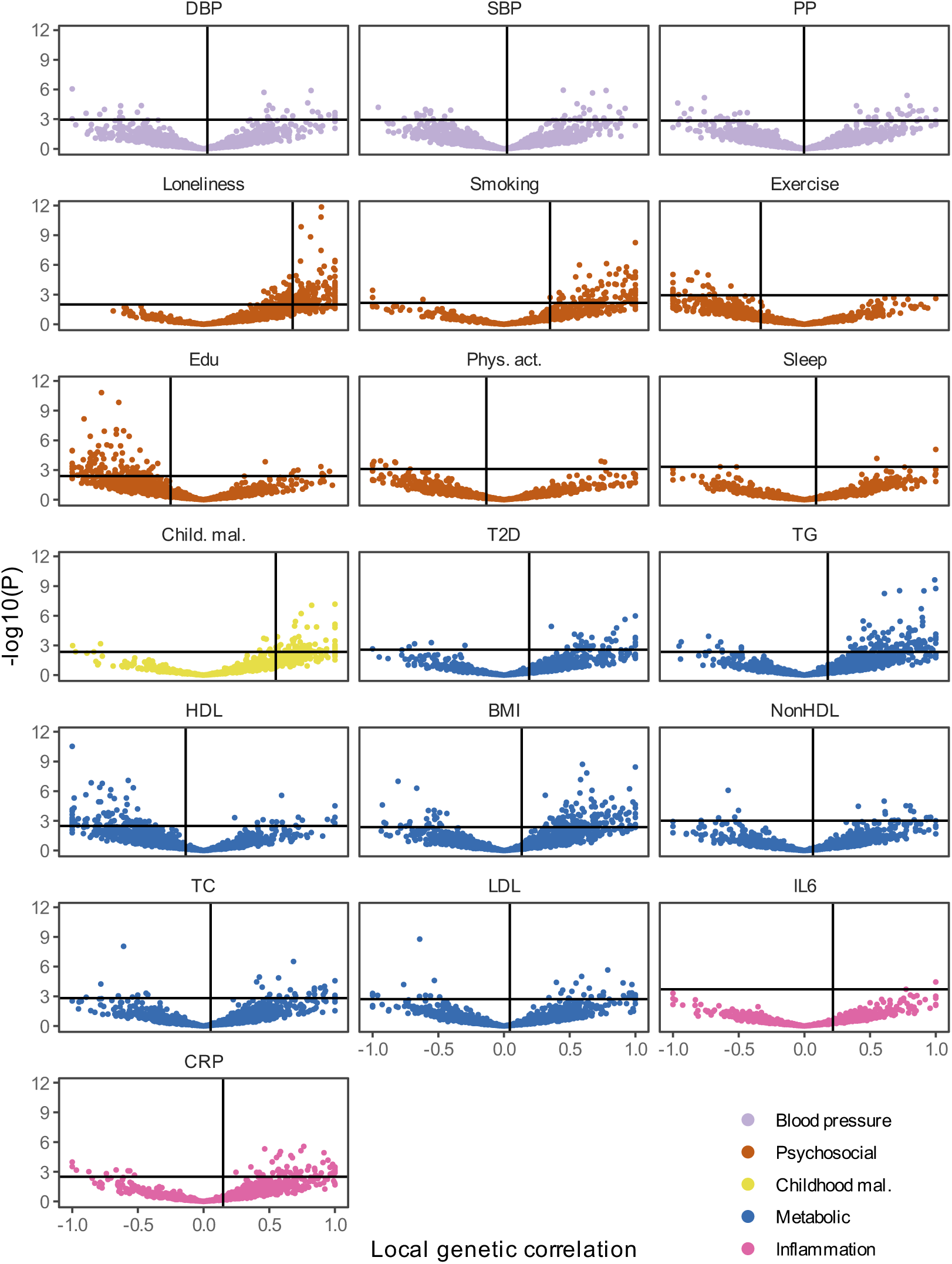
Local genetic correlations between MDD and risk factors. Volcano plots based on LAVA results. Local genetic correlation between MDD and each of the risk factors (x-axis) and the corresponding −log10 transformed *P-*value (y-axis). Correlations were estimated in the loci that showed marginally significant local heritability in 2,495 considered genomic regions. Loci exceeding the horizontal line are significant at *P*_FDR_<.05. Multiple testing was adjusted for individually for each trait over considered loci. Psychosocial=Psychosocial/Lifestyle; DBP=Diastolic Blood Pressure; SBP=Systolic Blood Pressure; PP=Pulse Pressure; Edu=Educational attainment; Phys. Act.=Physical activity; Child. Mal.=Childhood Maltreatment; T2D=Type II Diabetes; TG=Triglycerides; HDL=High-Density Lipoprotein; NonHDL=Non-High-Density Lipoprotein; TC=Total Cholesterol; LDL=Low-Density Lipoprotein; IL6=Interleukin-6; CRP=C-Reactive Protein

**Extended Data Fig. 8.**
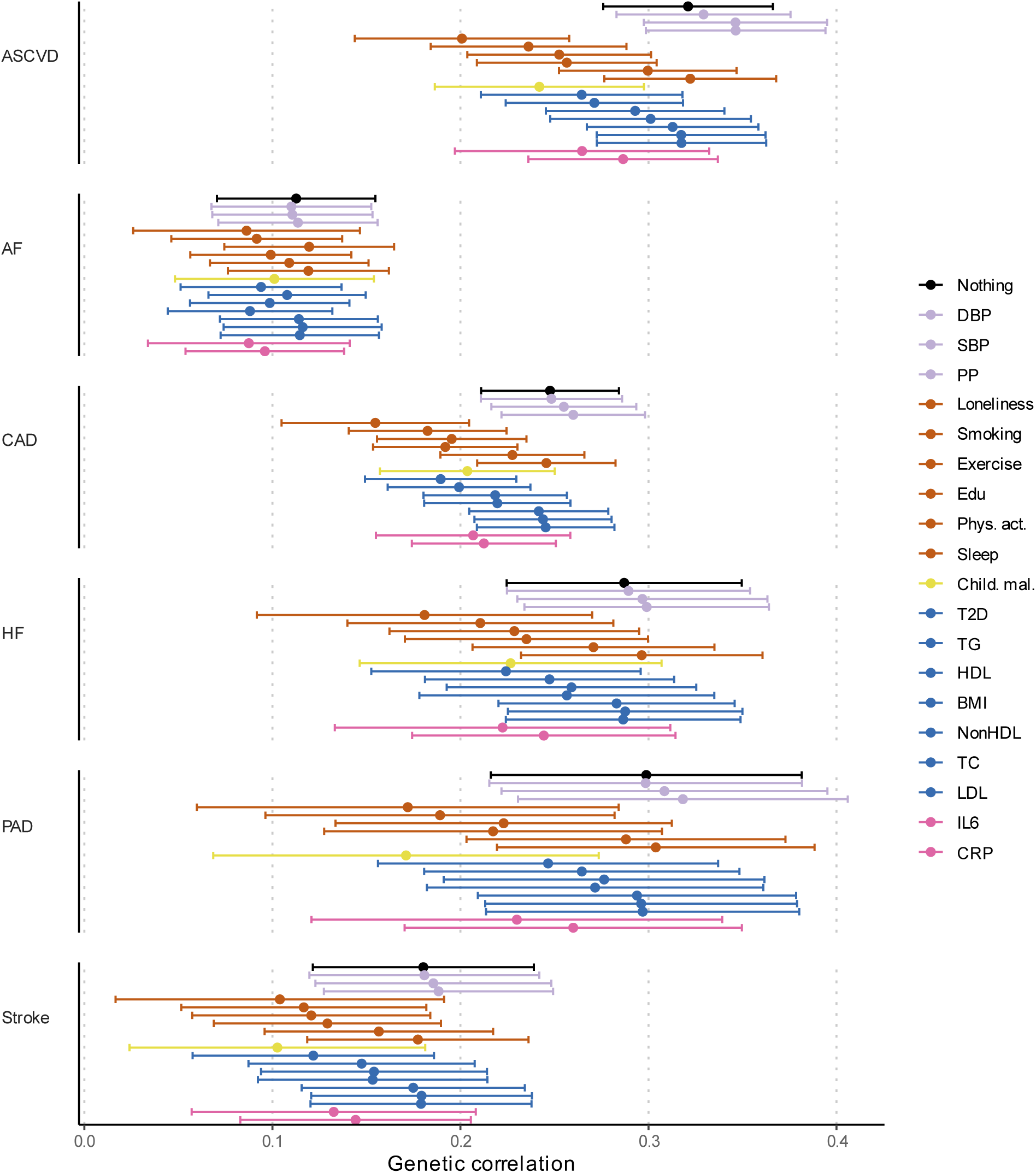
Genetic correlation between MDD and CVD when adjusting for individual risk factors. Results from Genomic SEM. Bars represent 95% confidence intervals. Reference estimates of the association without any adjustment are printed in black. AF=Atrial Fibrillation; CAD=Coronary Artery Disease; HF=Heart Failure; PAD=Peripheral Artery Disease; DBP=Diastolic Blood Pressure; SBP=Systolic Blood Pressure; PP=Pulse Pressure; Edu=Educational attainment; Phys. Act.=Physical activity; Child. Mal.=Childhood Maltreatment; T2D=Type II Diabetes; TG=Triglycerides; HDL=High-Density Lipoprotein; NonHDL=Non-High-Density Lipoprotein; TC=Total Cholesterol; LDL=Low-Density Lipoprotein; IL6=Interleukin-6; CRP=C-Reactive Protein

**Extended Data Fig. 9.**
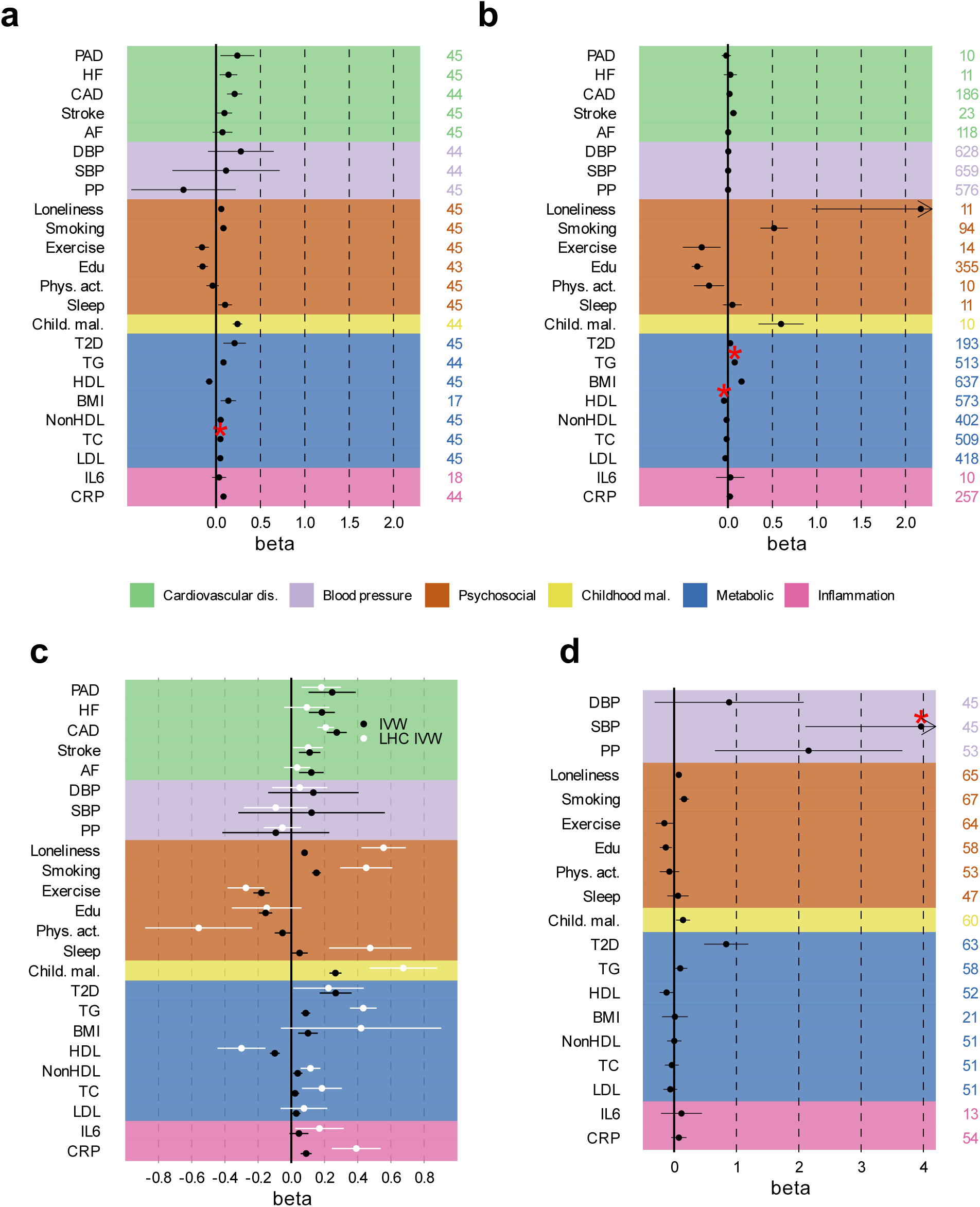
Assessment of evidence for causal associations between MDD, CVD, and risk factors. Results from univariable and multivariable Mendelian randomization (MR) analysis (Inverse variance weighted estimates are shown**) (a)** Effect of liability to MDD (exposure) on CVD and risk factors (outcomes) after excluding the UKB sample from MDD (which was responsible for most of the sample overlap in the exposure and outcome GWAS summary statistics). **(b)** Estimated fffect of CVD and risk factors on MDD after excluding the UKB. **(c)** Results from LHC-MR, that corrects for heritable confounders and sample overlap, with the standard inverse variance weighted MR estimate given as reference, for the effect of MDD on CVD and risk factors. **(d)** Effect of liability to MDD-ASCVD on outcomes and risk factors. **A-c** Statistically significant pleiotropic estimates are indicated with a red asterisk. The term beta refers to the log odds ratio. Psychosocial=Psychosocial/Lifestyle; AF=Atrial Fibrillation; CAD=Coronary Artery Disease; HF=Heart Failure; PAD=Peripheral Artery Disease; DBP=Diastolic Blood Pressure; SBP=Systolic Blood Pressure; PP=Pulse Pressure; Edu=Educational attainment; Phys. Act.=Physical activity; Child. Mal.=Childhood Maltreatment; T2D=Type II Diabetes; TG=Triglycerides; HDL=High-Density Lipoprotein; NonHDL=Non-High-Density Lipoprotein; TC=Total Cholesterol; LDL=Low-Density Lipoprotein; IL6=Interleukin-6; CRP=C-Reactive Protein

**Extended Data Fig. 10.**
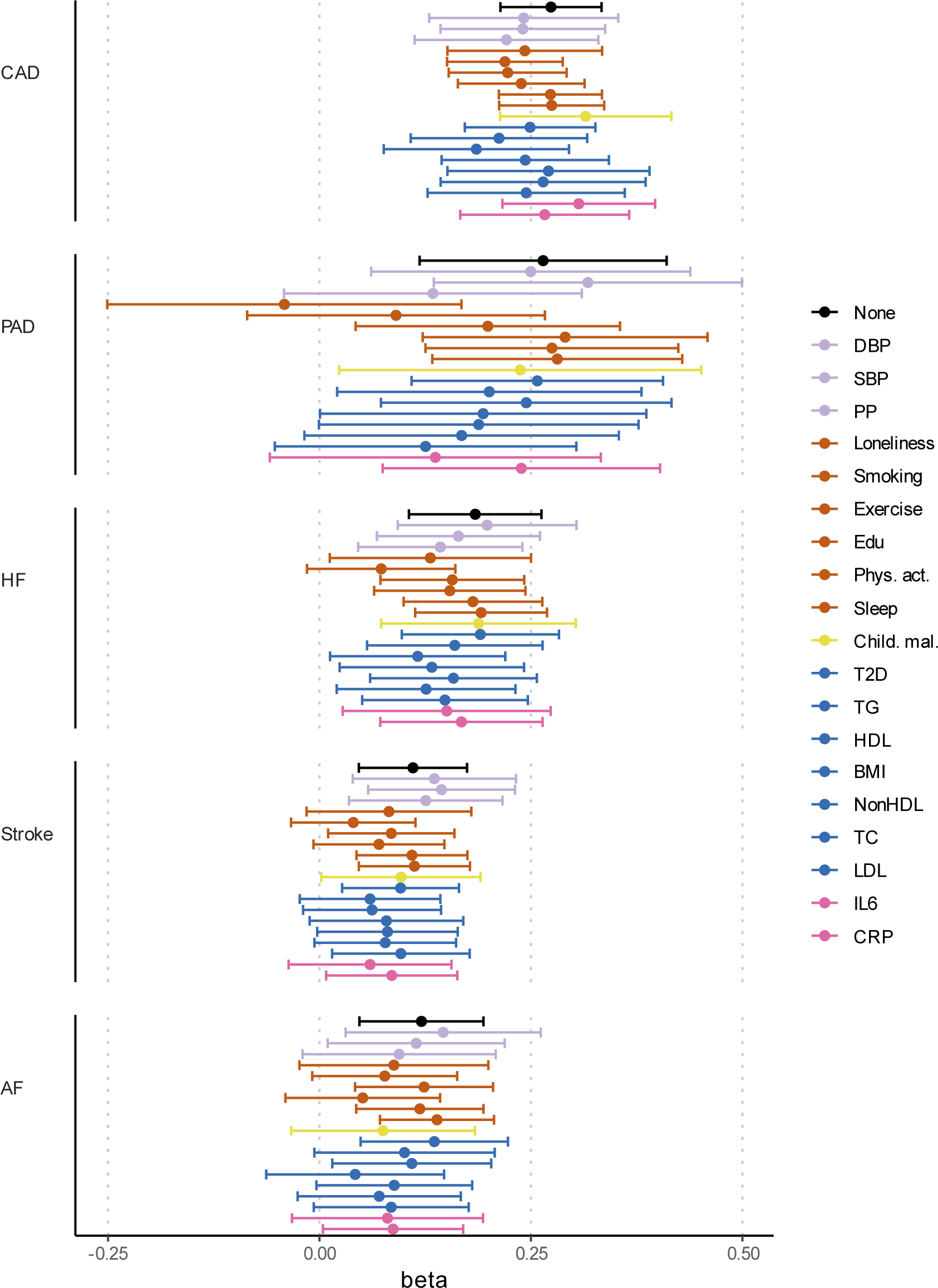
Effects of liability to MDD on CVDs when adjusting for individual risk factors. Results from multivariable Mendelian randomization. Bars represent 95% confidence intervals. Reference estimates of the association without any adjustment are printed in black. AF=Atrial Fibrillation; CAD=Coronary Artery Disease; HF=Heart Failure; PAD=Peripheral Artery Disease; DBP=Diastolic Blood Pressure; SBP=Systolic Blood Pressure; PP=Pulse Pressure; Edu=Educational attainment; Phys. Act.=Physical activity; Child. Mal.=Childhood Maltreatment; T2D=Type II Diabetes; TG=Triglycerides; HDL=High-Density Lipoprotein; NonHDL=Non-High-Density Lipoprotein; TC=Total Cholesterol; LDL=Low-Density Lipoprotein; IL6=Interleukin-6; CRP=C-Reactive Protein

